# What are the mechanisms of effect of group antenatal care? A systematic realist review and synthesis of the literature

**DOI:** 10.1101/2023.10.09.23296763

**Authors:** Anita Mehay, Giordana Da Motta, Louise Hunter, Juliet Rayment, Meg Wiggins, Penny Haora, Christine McCourt, Angela Harden

## Abstract

**Background:** There is growing interest in the benefits of group models of antenatal care. Although clinical reviews exist, there have been few reviews that focus on the mechanisms of effect of this model.

**Methods:** We conducted a realist review using a systematic approach incorporating all data types (including non-research and audiovisual media), with synthesis along Context-Intervention-Mechanism-Outcome (CIMO) configurations.

**Results:** A wide range of sources were identified, yielding 100 relevant sources in total (89 written and 11 audiovisual). Overall, there was no clear pattern of ‘what works for whom, in what circumstances’. Findings revealed six interlinking mechanisms, including: social support, peer learning, active participation in health, health education and satisfaction or engagement with care. A further, relatively under-developed theory related to impact on professional practice (but was relatively under-developed). An overarching mechanism of empowerment featured across most studies but there was variation in how this was collectively or individually conceptualised and applied.

**Conclusions:** Mechanisms of effect are amplified in contexts where inequalities in access and delivery of care exist, but poor reporting of populations and contexts limited fuller exploration. We recommend future studies provide detailed descriptions of the population groups involved and that they give full consideration to theoretical underpinnings and contextual factors.

**Registration:** The protocol for this realist review was registered in the International Prospective Register of Systematic Reviews (PROSPERO CRD42016036768)

## Background

Antenatal care (ANC) is considered an integral component of maternity care and can make a vital contribution to improving health outcomes and reducing inequalities [1]. According to the UK’s National Institute for Clinical Excellence (NICE), pregnancy care should be woman-centred and enable informed decision making [2]. Some women are well prepared for the challenges brought on by the journey through this major life event, but many experience significant barriers to enabling optimal care for themselves and their babies [3]. There is growing evidence to suggest that care should be tailored to meet the diverse needs of women and birthing people, within sometimes complex social situations [4] but evidence on its implementation is scarce [3]. Many women report being overwhelmed with so much information and that care is not person-centred, particularly in hospital settings [5]. Within the context of hospital maternity services, in many countries, antenatal care is fragmented, leaving women feeling like ‘a number’ [6] with screening tests dominating antenatal appointments [7]. For many people from socially and ethnically diverse groups, the political, policy, clinical and philosophical contexts of maternity services make engagement with care challenging[8], leading to poorer maternity outcomes [9, 10]. Pregnant women (and their partners) are sometimes offered antenatal education classes. However, access and provision is inequitable across the UK and many do not attend due to cost and/or other constraints. Classes are considered important for providing information and facilitating social support, which is known to be important for short- and long-term wellbeing [11]. However, there is no consistent evidence that standard (didactic) antenatal education improves birth and parenthood outcomes and/or experiences [12].

A recent UK enquiry into maternal and child health highlighted the significantly higher mortality rates among women and babies from minority ethnic groups and those affected by social or economic deprivation and identified sub-optimal care experiences as a contributing factor [10]. Systemic, structural, and institutional factors can produce these health disparities and expose a pattern whereby women from socially and ethnically diverse groups receive inadequate maternity care. Frequently, there are multiple forms of intersecting inequalities which compound and create challenges and disadvantages based on numerous factors [13].

### Existing evidence on group antenatal care

Ensuring quality, equitable maternity care requires the development and evaluation of new care models and, where appropriate, scale-up and replication for maximum population health impact. Quality maternity care must incorporate medical checks, effective health information sharing. social support, and cultural safety for all women, to enable participation in timely and comprehensive care seeking. Satisfying and optimal care and outcomes may be supported with such holistic ANC models.

Group ANC is a care model combining elements of clinical assessment and learning with the aim of facilitating social connections [14]. One of the most established models is ‘Centering Pregnancy’, developed by a midwife in the US to tailor care to the needs of socially disadvantaged communities who experience poorer access and care quality [15]. Centering Pregnancy combines clinical checks with information sharing and is typically provided by the same two midwives facilitating a group of around 8-10 pregnant women. Individual clinical checks (palpations) are brief and conducted privately within the same space as the group. The model was developed in response to recognition of the importance of social support during pregnancy and the transition to parenthood, and known limitations of didactic approaches to teaching and learning. Furthermore, women are not viewed as passive recipients of care, but are encouraged to make informed decisions, provide informed consent (or refusal), and to take an active role in their care to attain the best outcomes for themselves and their babies.

A 2015 Cochrane review of experimental studies concluded there is not yet sufficient evidence to draw strong inferences about clinical outcomes [16] and a later systematic review of randomised controlled trials and cohort studies did not find significant differences in clinical outcomes [17]. However, a review focused on outcomes for women categorised as higher risk showed more variable effects, with greater benefits for specific groups including adolescents and African American women [18]. Group ANC is a complex, person-centred intervention, therefore it cannot be assumed that benefits identified in one study or setting will be scaled and replicated in others [19]; evaluations need to take account of practice variations and local contexts, including beliefs and views of local health professionals and of service users [20]. Group ANC also combines different components (i.e., continuity of midwifery carer, social support and enquiry-based learning) which in themselves may have different explanatory theories of effect. Emerging evidence suggests that empowerment and support are core principles of group ANC, which yields benefits for women in contexts with inequalities in access and delivery of care [21]. However, there are different theoretical perspectives to understanding the mechanisms of empowerment [22] and within group ANC, the concept is still under-theorised and poorly understood. Increasing our analytical understanding of the theoretical propositions that underpin group ANC, the ‘ingredients’ of the model, will help to explain any effects and the role of context, to support further developments of the model and inform scaleup and replication/adaptation. This calls for an approach rooted in critical realism [23] to better understand the underlying causal mechanisms and the interplay between observable and hidden mechanisms shaping how group ANC might work for particular groups and within different contexts.

### Realist synthesis approach

Realist synthesis is an approach to systematic review and synthesis which focuses on identifying and testing potential context-intervention-mechanism-outcome configurations to develop theoretical and substantive understanding of how an intervention works, for whom and in what circumstances [24, 25]. It challenges positivist models of systematic review by positing that complex interventions do not ‘work’ in an ‘a-contextual’ and standardised fashion, replicable once subjected to rigorous evaluation. Instead, mechanisms of effect are produced by the ways in which interventions are interpreted, implemented and enacted, in particular environments and by people who may actively shape them [26]. In order to develop an appropriate experimental study, therefore, we identified a need to clearly understand potential mechanisms of effect of this care model and to develop a context-sensitive model which includes a core set of components around which local implementation would vary [27]. In this sense, realist reviews seek to provide explanations rather than measure outcomes.

This review was developed as part of a broader research programme, the REACH Pregnancy Programme [28], which sought to develop, implement and evaluate a bespoke model of group ANC (called ‘Pregnancy Circles’) for a socially and ethnically ‘superdiverse’ community [29]. The primary aim of this realist review was to articulate both implicit and explicit theories of action and key principles of group ANC. Secondary aims were to synthesise the findings/methods of the sources under review in relation to maternal and newborn health and wellbeing, and health services/service provider outcomes. The specific objectives were to:

1. Identify and review relevant research on/reports of implementation of group ANC models.
2. Articulate theories informing the models evaluated.
3. Identify the context and mechanisms of change in models already evaluated, recognising the likely complexity.
4. Synthesise and develop a set of core principles to inform the design and development of an intervention model tailored for our context named ‘Pregnancy Circles’.
5. Inform the preparations for implementing and testing the model in a planned multi-centre RCT.
6. Synthesise the findings of the range of the studies/sources on the subject.

## Methods

This review was conducted following the RAMESES guidelines for realist synthesis, and the PRISMA guidelines for systematic reviews [30, 31].

### Eligibility criteria

We envisioned different sources would contribute different context, intervention, mechanisms and outcome (‘CIMO’) insights (with some containing several data types). We therefore sought to mine for theoretical and empirical data in a wide range of media, including clinical trials, qualitative studies, reviews, reports, commentaries and videos. We included sources describing reviews as background information to provide theoretical insights; only sources describing primary research were accessed for data extraction and analysis. Non-research sources (e.g. opinion pieces, audio-visuals) were also included as these may highlight theoretical propositions underlying model development and implementation.

Inclusion criteria:

1. All sources related to any outcomes of an ANC model, or ANC and postnatal care that include participants meeting in a group (at least 4 women)
2. All sources related to the process or implementation of an ANC model that includes women meeting in a group (more than 4 women)
3. All sources related to experiences of an ANC model that includes women meeting in a group (>4 women) (professionals’ or users’ experiences)
4. All national/country contexts

Exclusion criteria:

i. Groups that do not include ANC
ii. Groups provided outside NHS/mainstream health care (e.g., by charity groups)
iii. Groups that provide speciality rather than routine care (e.g., obesity ‘clinic’)
iv. One-off groups
v. Groups not including clinical care (e.g., classes only)
vi. Groups not involving any health professional input (e.g., peer-led groups)

Following data extraction, a further exclusion was applied prior to analysis:

vii. Sources relating to opinions and experiences without relevant CIMO data.

### Study selection

Database searches were conducted in MEDLINE, PsychINFO, EMbase, Maternity and Infant Care, Web of Science, Cochrane library (Cochrane Central Register of Controlled Trials [CENTRAL] and Database of Systematic Reviews) (see Appendix 1 for search terms used). No language restrictions were imposed for the initial search. Sources published from and including January 1980 to March 2015 were eligible for inclusion. Grey literature was sought in sources including OpenGrey, GreySource, internal reports and non-peer reviewed journals such as Midwifery Digest). Reference chaining was undertaken on all relevant sources retrieved, and forward and back-citation searches conducted using Google Scholar. Searches were also undertaken in relevant websites such as the Centering Healthcare Institute Inc., Association for Improvements in Maternity Services, National Childbirth Trust, and Local Supervising Authority Midwifery Officers Forum. As we aimed to include audio visual media, we also searched YouTube and internet search engines using key terms.

EPPI-Reviewer version 4 was utilised for data/review management. Titles and abstracts of written sources retrieved were first independently double screened by two researchers with any differences resolved through discussion or deferred to full text assessment. Full texts of included written sources were then double screened by two researchers, and any disagreements adjudicated by a third person. Audio visual media sources were screened by one reviewer using the same criteria. Realist reviewing is complex and time-consuming so following the analysis, the search was updated in April 2019 to identify any additional sources or insights. These were screened and read analytically by two reviewers to identify whether any new themes should be added, or existing themes modified in light of new literature.

### Data extraction and management

We developed data extraction proformas to draw out data (i.e., descriptive notes, ideas and annotations or excerpts) around the terms of what works, for whom, in what circumstances. The extracted data were then interrogated by sub-teams of researchers to answer the research aims specifically relating to:

- **What works, and for whom?** Outcomes measured in each study were collated and compared by study population, to determine whether they were more or less successful with different groups of women (for example, vulnerable, young or socio-economically deprived groups).
- **How?** Are any explicit theoretical claims made about how the intervention might or did achieve the intended or experienced outcomes? What can be gathered implicitly regarding theories of how the intervention might or did work?
- **In what circumstances?** How does context at a strategic, institutional, inter-personal and individual level disrupt or support the implementation or delivery of the intervention?

The analysis was conducted inductively, with no initial attempt to impose a preconceived framework, theory or theories onto the data unless deemed appropriate. As part of the reflective process team members formulated their own logic models prior to analysis, to make their own ‘theories’ explicit; these were set aside for later reflection on the findings rather than used as a framework for analysis. Critical discussions were held within and between sub-teams during this process, and the data relevant to each question were then synthesised.

### Assessment of risk of bias

Reviewers assessed and ranked source quality and relevance and provided rationales for their decisions drawing on the RAMESES Quality Standards for Realist Synthesis [30]. Key principles by which sources were assessed included: whether they contributed to the development or testing of programme theories; and rigour: whether the research sources used credible and trustworthy methods. We used an adapted checklist from the Critical Appraisal Skills Programme (CASP) to assess rigour of research-based sources. Additionally, an overall assessment rating of low, medium or high was assigned, relating to the source’s usefulness for the review with ‘high’ rated sources prioritised during data synthesis.

### Data synthesis

We used a two-stage approach to synthesis; an initial analysis identified themes from the review data on what works for whom, how, and in what circumstances. A second, narrative synthesis, iteratively developed overarching themes through data interrogation and review team discussions. Statistical analysis based on any group or sub-group outcomes was outside the review scope and focus.

## Results

### Identification, screening and study selection

*The initial electronic search in 2015 produced 2,238 records with another seven sources obtained from hand searching of reference lists. After full screening, the review included 100 sources (of which 11 were audiovisual and the remaining 89 written sources) (see* Figure 1). An updated search in April 2019 identified 75 additional sources, of which 48 met the inclusion criteria and 27 were excluded following full text review (two of which were study team publications). Of those 48 sources, 15 were conference abstracts or posters which did not provide sufficient detail to add new insights to the analysis. Of the remaining 33 sources, there were no additional or divergent themes identified and they were therefore not included in this synthesis (summary details are given in Supplementary file 1 and 2). As a result, we concluded that analytical saturation had been reached and no further search updates were conducted.

**Figure 1:**
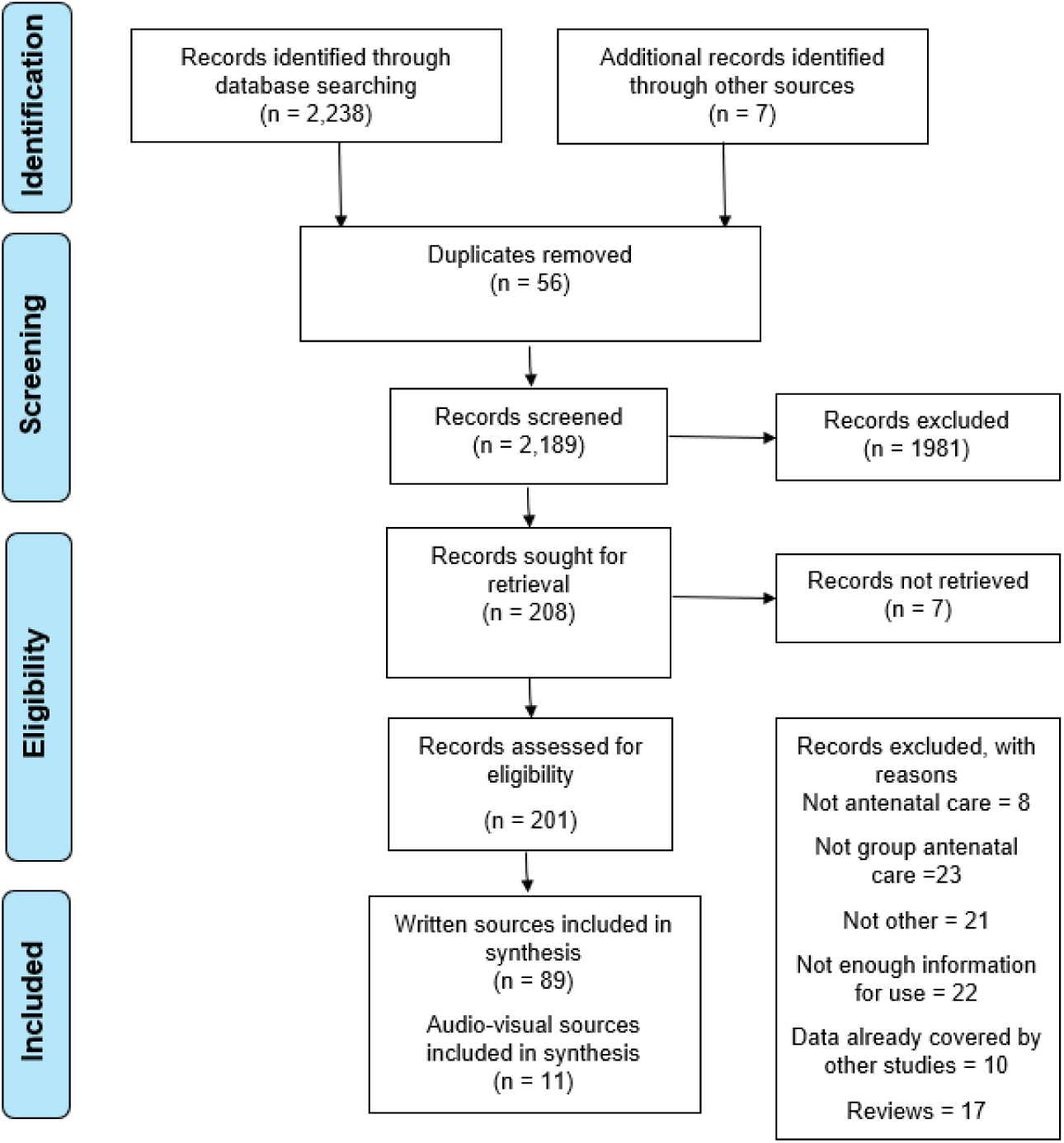
Consort diagram.

### Characteristics of included sources

Most sources were either cohort studies or opinion/expert reviews. Only four sources were from the Global South. Most sources (n=77) related to populations and contexts in the United States. There were 14 studies or projects which had numerous sources reporting on outcomes or were commentaries, editorials or conference abstracts related to that study. The sources from the same project or study were linked for the review and data extraction to avoid data duplication.

The vast majority referred to a ‘standard’ CenteringPregnancy (n=62). This follows the recommended schedule of ANC visits (lasting 90 minutes to two hours long) with women taking and recording their own health data, facilitative discussions, and activities to address important health topics, with private time with their provider. Of the CenteringPregnancy interventions, six described the model with ‘additions’ (including dental oral health components or specific topics relating to adolescents and youth). Another six sources described CenteringPregnancy with ‘adaptations’ where the private provider time was either scheduled outside of the main group session (i.e., either side of it), or where ANC visits involved a combination of one-to-one appointments and group sessions throughout pregnancy. There were seven sources describing non-CenteringPregnancy models of group ANC, which broadly described similar models of care to standard CenteringPregnancy.

### Findings

#### What works and for whom?

Evidence on the benefits for particular population groups was inconsistent when examining ‘what works’. We coded and categorised outcomes into four key domains: 1) experience (e.g., satisfaction), 2) clinical (e.g., mode of birth, birth weight), 3) health behaviours (i.e. smoking, breastfeeding), and 4) psychosocial (e.g. self-efficacy). We then coded population groups into 10 categories based on four population risk factors (social/demographic factors; medical, economic or none) within high income or low to middle income countries (see **Error! Reference source not found.**).

**Table 1:** Categories of population groups.

|  | High income country | Low-income country |
| --- | --- | --- |
| **General** | 1. General population with no reported ‘risks’<br>2. Unknown (not stated) | 3. General population with no reported ‘risks’ |
| **Economic risk factors** | 4. Low income | - |
| **Medical risk factors** | 5. Underserved<br>6. High-risk<br>7. Low-risk | - |
| **Social and/or demographic risk** | 8. Military groups<br>9. Minority ethnic groups | - |
| factors | 10. Young |  |

Mapping the outcome categories by population group categories generated no overall conclusive patterns as to what works for any particular population groups, although there were some indications of benefits for military families [32], for African American women [33] and for adolescent mothers [34].

Poor reporting and rationale for targeting particular population groups hampered comparisons. For example, some sources stated the targeted population groups were socially or medically high-risk but did not explain in detail or provide a clear rationale for why and how group ANC was expected to confer benefits. Others defined risk by the geography of an area such as where a clinic was located (i.e., low-income area) but did not explain this further in relation to the group care participants. Furthermore, being from a minority ethnic group was deemed high-risk due to the increased prevalence of poorer clinical outcomes at the population level, with little detailed understanding of how race and ethnicity were associated with poorer outcomes. For example, group ANC tended to have limited effectiveness compared with usual care in communities where women already had strong social support networks [35, 36]. Some sources also referred to the ‘Latina paradox’ whereby group ANC had little positive effect due to the already high levels of social support found within the Latin American population [36]. No studies examined the interconnected nature of multiple and compounding risk factors (i.e., through an intersectionality lens).

#### How (mechanisms of effect)?

Most sources drew on implicit rather than formal explicit theories to explain how group ANC might work to improve outcomes. Most sources also described CenteringPregnancy therefore they shared common theorised mechanisms of how group ANC might work. All implicit and explicit explanations were drawn out and coded, which generated six broad mechanisms of effect: 1) Social support; 2) Peer Learning; 3) Active Participation in Health; 4) Health Education; 5) Satisfaction with care; 6) Health Professional Development and Wellbeing (see **Error! Reference source not found.**).

**Table 2:**
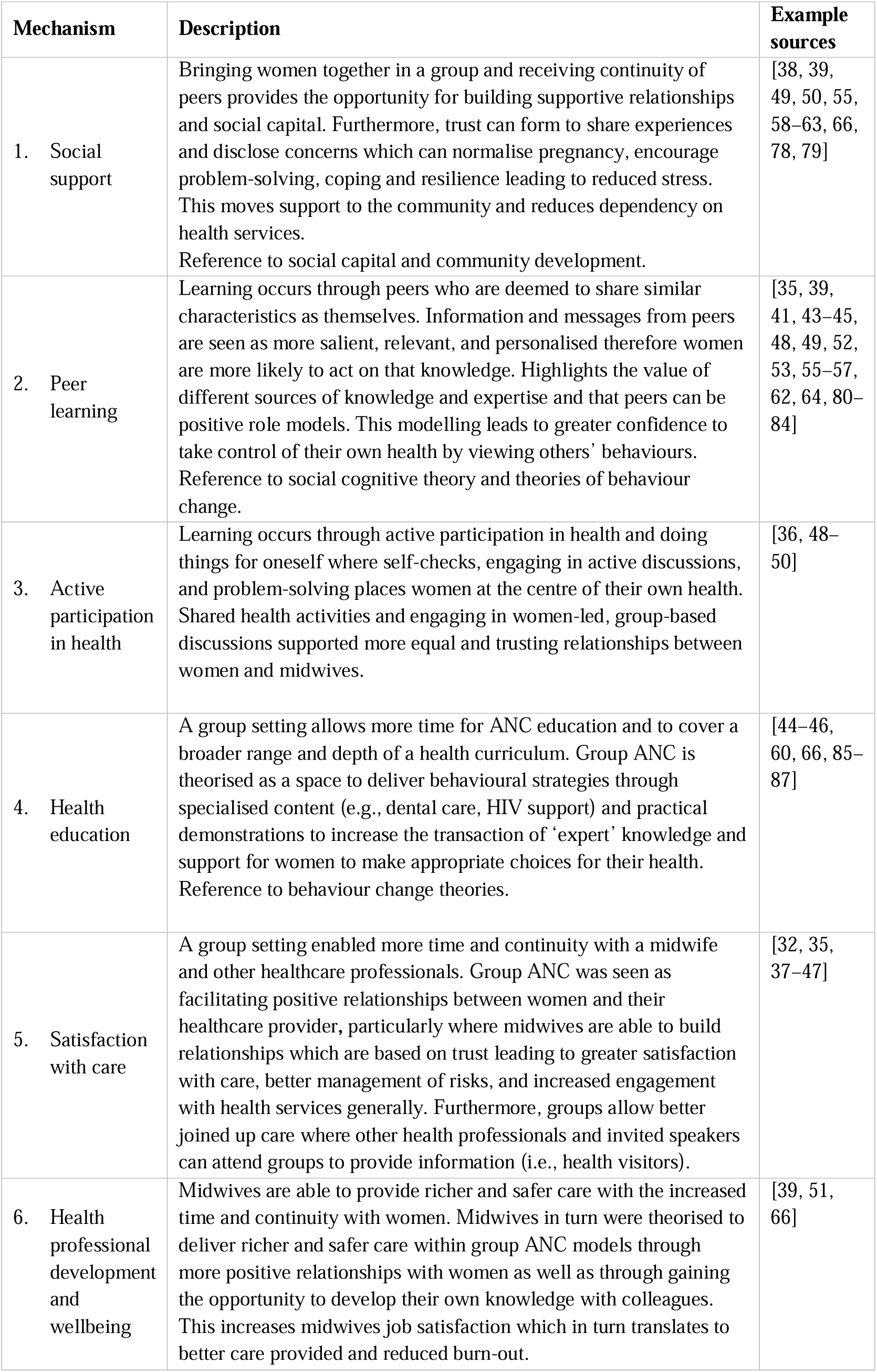
Theorised mechanisms of effect.

Most sources referred to a number of these mechanisms, but some focussed on one or two only; for example, transaction of knowledge and health persuasion messaging (an individualised theory) while others were instead focused on the exchange of peer knowledge and breaking down of traditional ‘expert knowledge’ sources (a collective theory of action).

Group ANC was believed to facilitate positive relationships between women and their healthcare provider where more time and continuity with midwives (and other health professionals) built additional trust leading to increased satisfaction and engagement with care, as well as management of risks that was more responsive [32, 35, 37–47]. Shared health activities and engaging in women-led, group-based discussions supported woman-midwife relationships that were more equal and trusting [36, 48–50]. Additionally, it was theorised that midwives deliver richer and safer care within group ANC models through more positive relationships with women and through gaining the opportunity to develop their own knowledge with colleagues [38, 51].

Sources referred to mechanisms relating specifically to the group element of care. For example, group modelling supported empowerment where women developed greater confidence to take control of their health by viewing others’ behaviours [43–45, 48, 49, 52–57]. It was theorised that group ANC provided peer and community support, allowing women to share and normalise experiences, whilst also gaining practical and relevant psychosocial support [37, 50, 58–63]. Reduced stress and increased coping skills through pregnancy were anticipated [64, 65]. It was expected that peer support would reduce unnecessary reliance on health services and build more resilient communities [35, 38, 39, 49, 55, 60, 66].

An overarching mechanism was the reoccurring concept of empowerment as related to increasing women’s knowledge, ability to make better informed decisions (and behaviour changes) and building positive support networks with healthcare providers, own peers and their communities. However, most sources poorly theorised the concept of empowerment with different underpinning assumptions from individualistic and collective perspectives. For instance, nearly all theorised a link between greater knowledge and empowerment, whereby active learning approaches (e.g., peer-led group discussions) results in more relevant and salient knowledge, leading to empowerment in decision making and positive behaviour changes. This process of empowerment was largely conceptualised through an individualistic lens relating to self-efficacy and control over one’s health rather than broader concepts of empowerment which instead encompass collective and/or group levels of empowerment and/or paradigmatic shifts in care delivery. However, there was insufficient detail in the data to enable a fuller exploration of how such differences in the hypothesised underpinning mechanisms may influence the implementation, process or outcomes of group care implemented in different settings.

#### In what circumstances?

There were three main context factors related to implementation and delivery of group ANC models. Factors included: 1) Focus on the community and hyper-local level; 2) Shifting care out of hospitals, 3) Adapting to a different way of working.

##### Focus on the community and hyper-local level

Most group ANC models of care sought to focus on the community and hyper-local level (i.e., particular populations and areas of deprivation) to signify equal partnership between women and facilitators [67]. Group ANC models needed to be easily accessible at this level to recruit and reach women whilst also working at a scale to allow for an appropriate number to form a group size of 8-10. This posed a number of practical challenges for teams, mainly recruiting a desired number of women at similar gestations within local areas which required good targeting, scheduling and organisation [37, 58]. Focused recruitment strategies were needed to encourage women’s interest and engagement e.g., vouchers, automated reminders and involving local women in setting up and promoting groups [50, 68]. An opt-out recruitment (rather than opt-in) was also used with success in another study [58]. There was some evidence that a lack of childcare facilities discouraged multiparous women from attending group care [36, 68]. Engaging women in early pregnancy was considered important (particularly if any behaviour change was a desired outcome) [43, 46, 69], however engaging women too early could lead to high discontinuation rates [41].

##### Shifting care outside hospitals

Most group ANC models were delivered in community settings rather than hospitals to aid accessibility, work at community and hyper-local level, and reach target populations. For example, an initiative targeting pregnant school pupils was held on a school site immediately after the end of the school day, which supported attendance [68, 70]. Sourcing suitable community venues, and the increased time taken to transport equipment and set up venues each week, was a recurring practical challenge [32, 37, 44, 66, 71]. Many community-based venues were often not immediately appropriate for medical tasks such as routine blood tests and accessing patient records [50, 51]. Group ANC required appropriate infrastructure and troubleshooting to manage the shift to care delivery outside of hospitals.

##### A different way of working

Groups were often set up and/or sustained by a small group of midwives or nurses who were committed to the concept, took ownership of the initiative and invested considerable time and effort to ensure its success [50]. The satisfaction gained from providing group ANC, working flexibly, making compromises where necessary and supporting each other enabled facilitators to sustain the model and overcome obstacles to implementation [58, 72]. Group ANC was usually a very new way of working for teams and services and there were key points of incompatibility to overcome. For example, group facilitation was not a well-developed skill for most midwives [73–75]. Adequate facilitator training was essential; and in its absence, both midwifery and medical professionals tended to adopt an overly didactic style, which was unsatisfying to women, leaving them feeling their concerns had not been addressed [40, 41, 76]. A didactic teaching style was deeply ingrained in some physicians [44], as reported in some Swedish and Canadian studies [41]. Women also needed to have a good understanding of the model otherwise there were confusions. For example, in one study, participants were not aware that group attendance replaced standard individual appointments and women tried to attend both [32]. Group ANC models also needed to consider the value this model offered within existing care. In one study, group ANC was less well received when set against a case-loading approach [66]. There was insufficient detail on national and local policies and health system factors to enable a full exploration of these broader context issues.

## Discussion

Overall, there was no clear pattern of ‘what works for whom, in what circumstances’. Variation in contexts, group ANC application or interpretation as well as which communities were involved may account for the inconsistency of findings. Our analysis did reveal some other important insights. We identified six interlinking mechanisms drawn out from mainly implicit descriptions. Mechanisms included: social support, peer learning, active participation in health, health education and satisfaction or engagement with care. A further theory related to impact on professional practice but was relatively under-developed. An overarching mechanism of empowerment featured across most studies, but most models largely adopted an individualistic lens despite the group/community focused approach. For example, some focused on the role of education, peers, and social support to change actions and behaviours. Others focused on broader paradigmatic shifts in professional-client relationships and the redistribution of power to women and communities. Conceptualisation of educational mechanisms drew on two somewhat different areas of pedagogical theory: one more focused on the emancipatory potential of the group approach to information and learning, whereas the other was rooted in a more transactional concept of education. The mechanisms of effect relating to empowerment were particularly important when considering which population group(s) to target for group ANC. There was poor reporting of populations, inadequate rationales for why particular populations were targeted and how the model was expected to confer benefits. For example, limited benefits were reported where women already had strong social support networks. No studies considered intersectionality of multiple and compounding risk factors. Few studies considered wider health system factors in shaping contexts, mechanisms and outcomes and most focused on site-specific context factors relating to implementation and individual/team level cultures. Much of the early conceptualisation and implementation of group ANC took place in the US, and it is possible that the model may function differently and have varying effects in different health systems, rather than simply in different local contexts or working with different populations and communities.

Our findings are largely in line with other research, including a previous Cochrane review demonstrating that there is insufficient evidence of benefits from this care model [16, 17]. We suggest there are inconsistencies in the evidence base due to variation in contexts, how the model is applied or interpreted as well as which communities are involved. For instance, our findings support other reviews which suggest that group ANC is likely to be most beneficial in groups and contexts with high levels of inequalities in access and experience of care, such as higher-risk or more vulnerable populations such as African American women and adolescents [18]. Another review focused on attributes that may support acceptability and effectiveness in LMICs and posited a generic model which was concordant with the mechanisms of effect we identified here, including empowerment and social support [21]. Since mechanisms of effect may have particular advantages in contexts where access and care inequalities exist, giving ‘women a voice for knowledge sharing and a sense of community support’ [21] may be of particular value.

### Strengths and weaknesses of the review

A realist approach helped to identify the potential mechanisms of effect for how group ANC ‘works’ with calls for more theoretical understanding about the concept of empowerment and how this relates to particular groups facing intersecting forms of inequality, disadvantage and discrimination. This approach also helped to provide more nuanced guidance on what to consider when implementing group ANC, including what features of the context are important. Our analysis however was limited by the lack of detail in study reporting, which meant some implicit understanding and insights had to be drawn out. Potential for researcher biases were handled through discussions and reviewing our own assumptions at review commencement to check how these may influence findings. In anticipation, each review team member drew a logic model at the outset, setting out their own theoretical propositions and assumptions about mechanisms of effect. Searches took place initially to feed into a feasibility study and development work for a bespoke model of group care to function in a UK NHS setting. A subsequent update identified no new themes relating to theories or mechanisms of effect. The team concluded that sufficient saturation was reached in the literature to inform future work to implement and evaluate this model of care (see Figure 2 and Figure 3).

**Figure 2:**
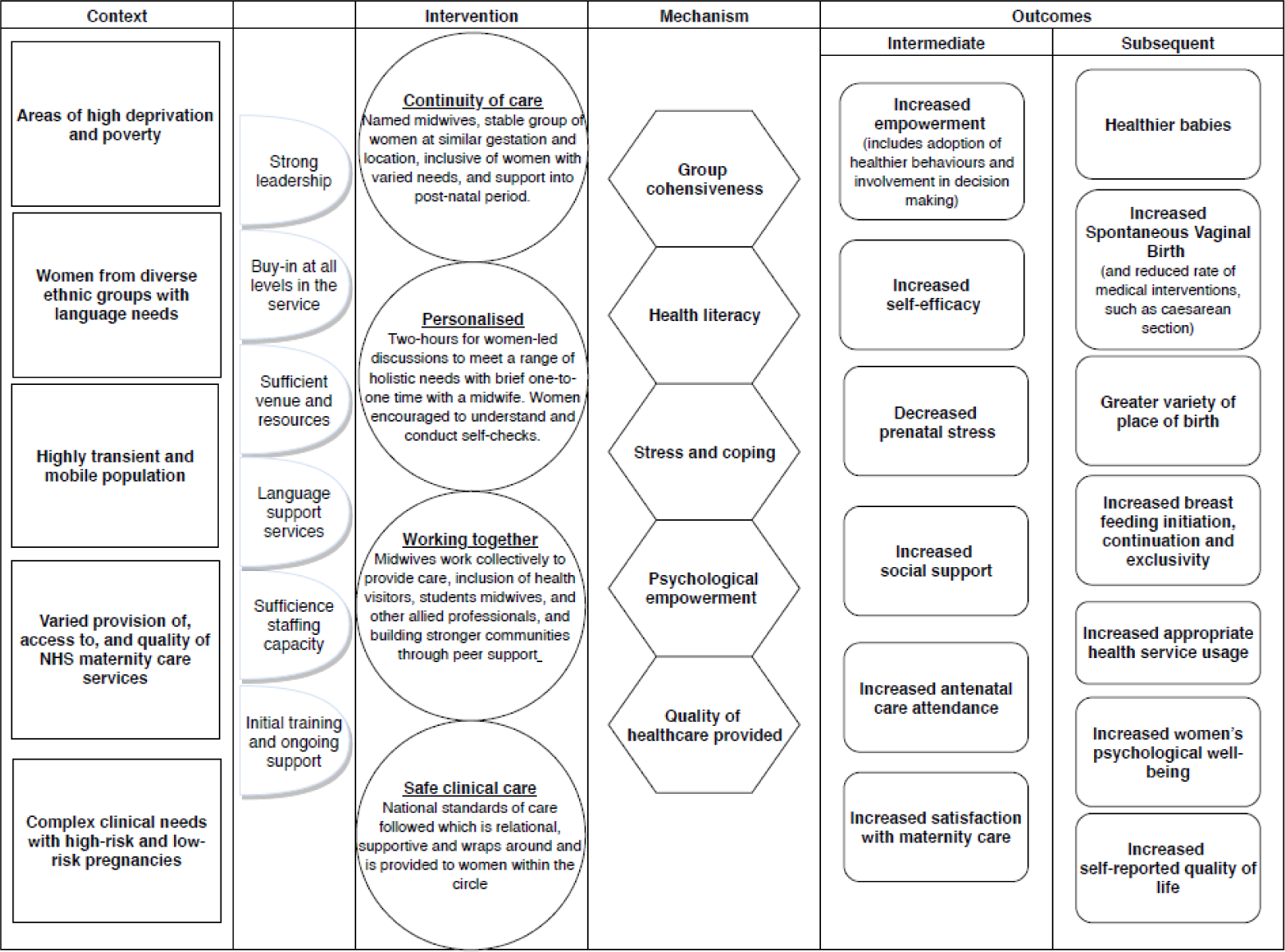
Pregnancy Circles logic model.

**Figure 3:**
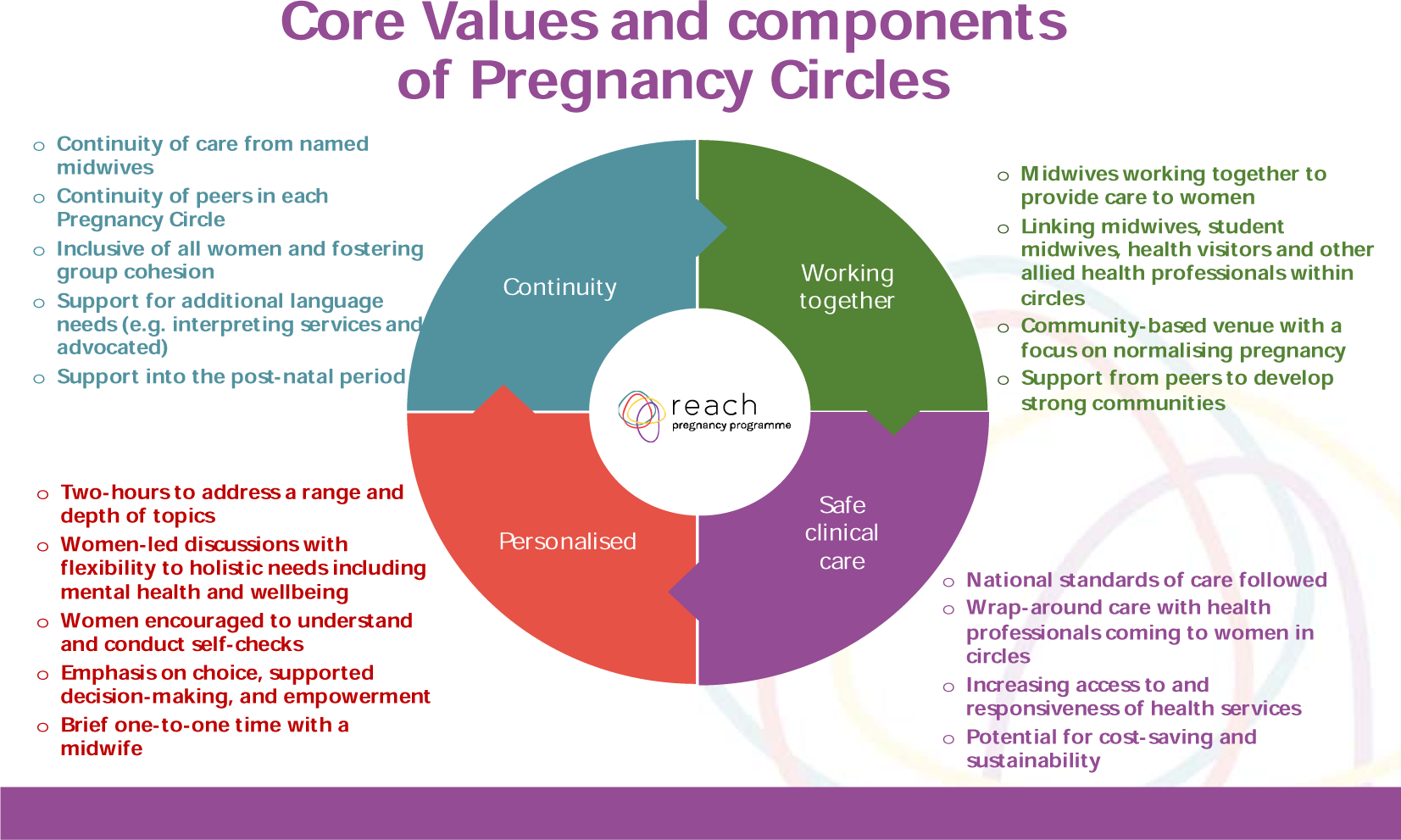
Pregnancy Circles values and components.

### Implications for researchers, care providers, and policymakers

This review of group ANC provides important implications for researchers, care providers and policymakers. Our review was hampered by the lack of study details particularly intervention and contextual descriptions and rationales for population group targeting. There is also a need for greater reporting quality and consistency. Future research would benefit from a clearer focus on mechanisms of effect, to ensure appropriate outcome measures are used, but also a clearer focus on who is expected to benefit and in which contexts. Further trials are essential, including detailed process evaluations exploring the role of care context and process, professional preparation and attitudes, the communities involved and how they experience group care. We identified few studies or other sources that examined the effects on care providers, and could not identify theories of how any impact on professionals may have an impact on service users. Much of the early conceptualisation and implementation of group ANC has taken place in the US, where the health financing system is not universal, access to healthcare is inequitable and midwives remain relatively marginal rather than mainstream healthcare providers. It is possible that the model may function differently and have varying effects in different health systems, rather than simply in different local contexts or working with different populations and communities. These should be considered in more depth in future studies and reviews. This review was undertaken alongside the conduct of a qualitative feasibility study and pilot trial. Both were intended to contribute to the conceptualisation and design of a contextually adapted model to be tested formally in a trial, with integral process evaluation. The findings of both studies were utilised to develop a logic and core values model for the trial intervention (see Figure 2 and Figure 3). An RCT with nested qualitative evaluation is currently in progress [28], following a successful pilot trial [77].

For providers and policymakers, we outline some key context factors which suggest a focus on supporting staff and teams to implement group ANC at a hyper-local community level and enabling the systems, infrastructure, time and training to shift care out of hospital settings and bring on broader paradigmatic shifts in care delivery and the women-provider relationship. Group ANC facilitators required support and learning to deliver the model in non-didactic ways and to bring out the benefits of the group dynamic. Further work is needed to examine the concept of empowerment, whether and how this may operate as an overarching mechanism of effect and in what circumstances.

## Data Availability

All data produced in the present work are contained in the manuscript

## Declarations

### Ethics approval and consent

Not applicable

### Consent for publication

Not applicable

### Availability of data and materials

All data generated or analysed during this study are included in this published article (and its supplementary information files)

### Authors contributions

AH is the PI for the project, and led the conceptualisation process with input from MW, CM, and PH. AM, GDM, LH, JR, MW, CM, and AH were involved in the data extraction and synthesis stages of the review. AM led the overall synthesis of findings with input from AH, CM, GDM. All authors read and approved the final manuscript.

## Acknowledgements

We would like to thank the EPPI-Reviewer team at the EPPI-Centre for their training and support and Silvia Potter for undertaking the initial searches and providing ongoing support.

## Appendices

### Appendix 1: Search terms

1. ANC terms

Descriptor, keyword, subject: childbirth or Prenatal Care OR Prenatal Diagnosis OR Perinatal Care or Maternal Health Services OR Obstetrical Nursing or parent education or parent education program or mothers education or fathers education or Fathers [education] or Mothers [education]

Tiab: pre-natal or prenatal or peri-natal or perinatal or ante-natal or antenatal or childbirth or parturition or obstetr* or neonatal or neo-natal or midwife or midwives or matern* or antepartum or ante-partum or peripartum or peri-partum

AND

2. Group care terms

Subject: Group processes or group process

Tiab: care model* or model* of care or model* of antenatal care or model* of prenatal care or model* of ante-natal care or model* of pre-natal care or circle* adj2 (education or class or classes or screening* or assessment* or checkup* or check-up* or check up*) or Group education or group class* or group screening* or group assessment* or group checkup* or group check-up* or group check up* or Group Family Nurse Partnership* or gFNP

OR

Search 2

Tiab: CenteringPregnancy or Centering Pregnancy or (group antenatal or group prenatal or group ante-natal or group pre-natal) adj1 (care or education or class* or assessment* or checkup* or check-up* or check up*)

Date range: 1980

## Supplementary files

**Supplementary file 1:**
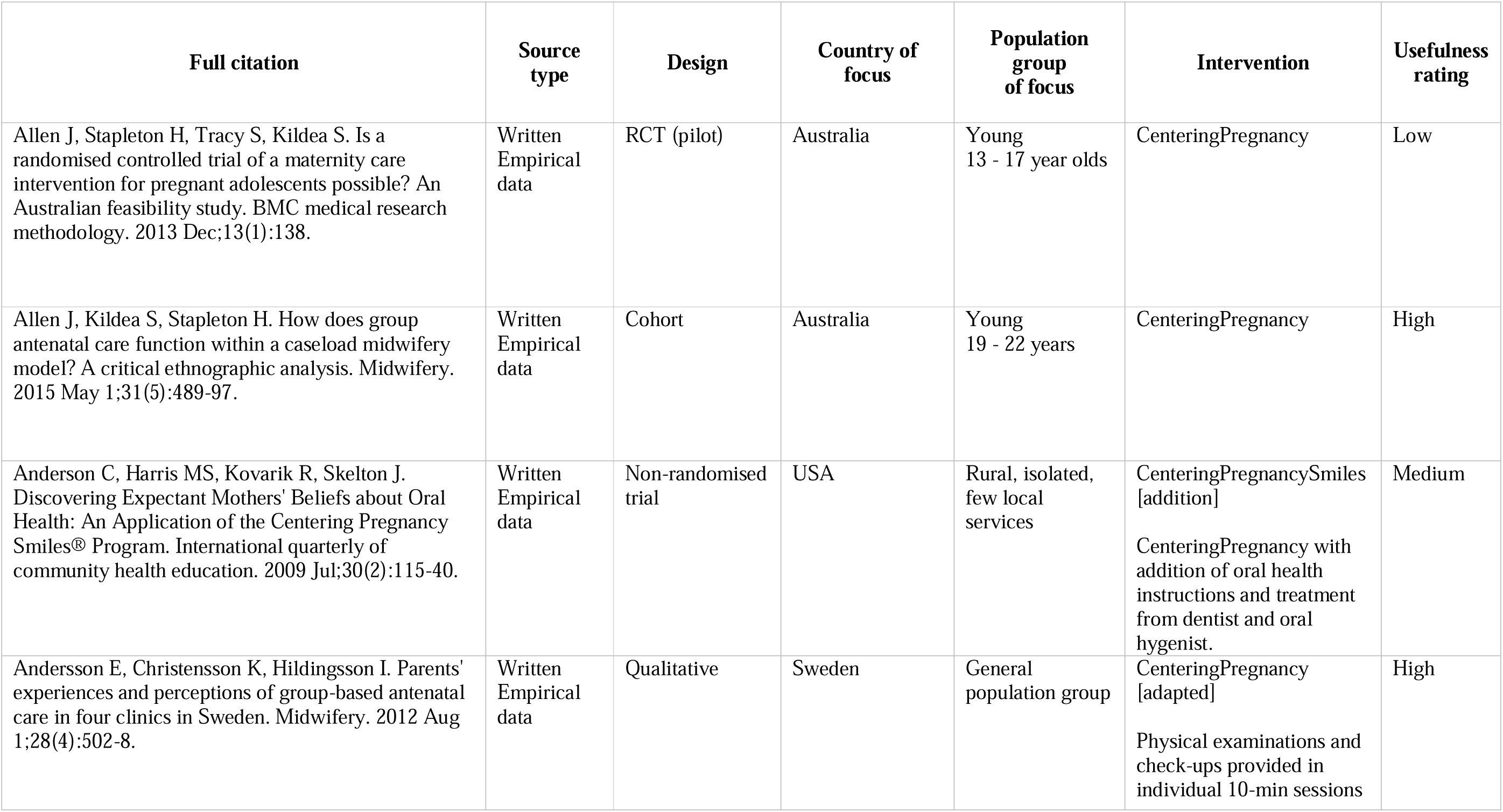
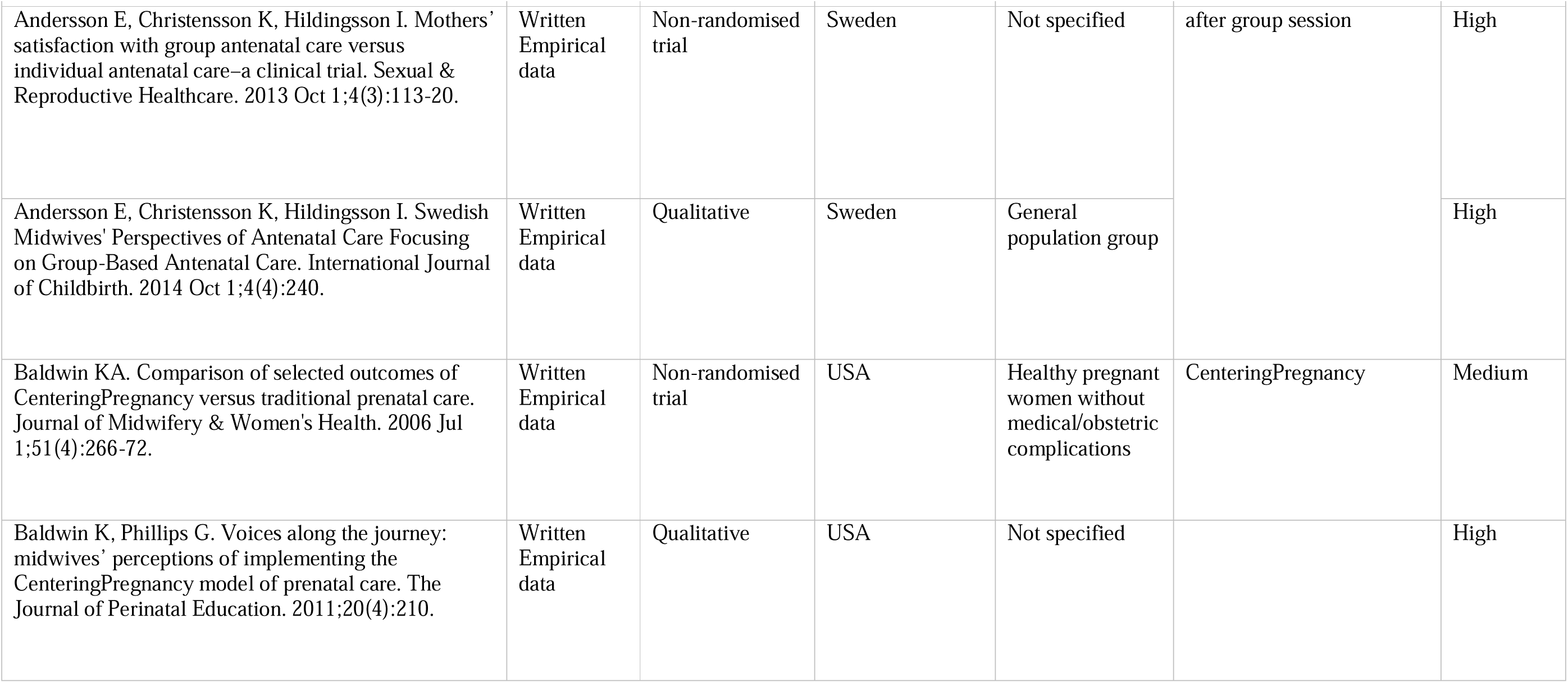
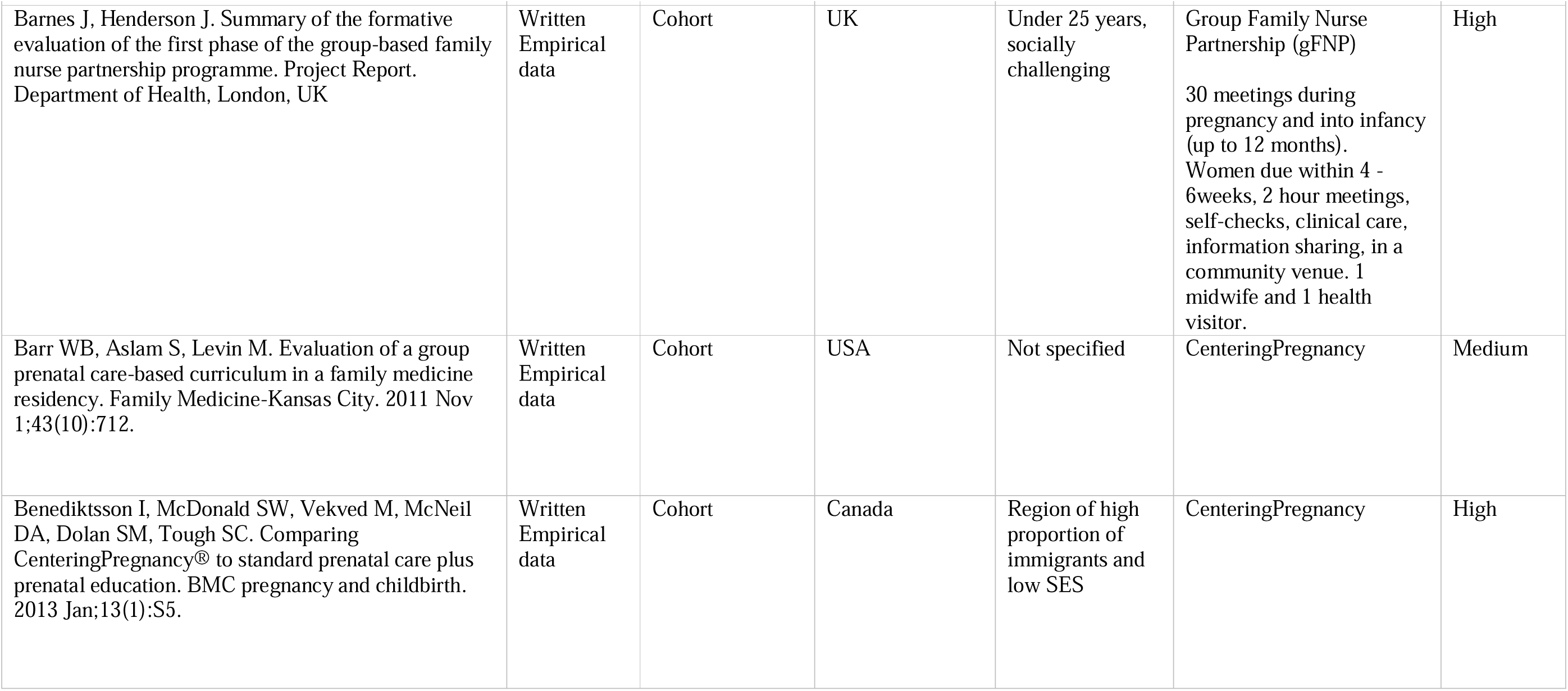
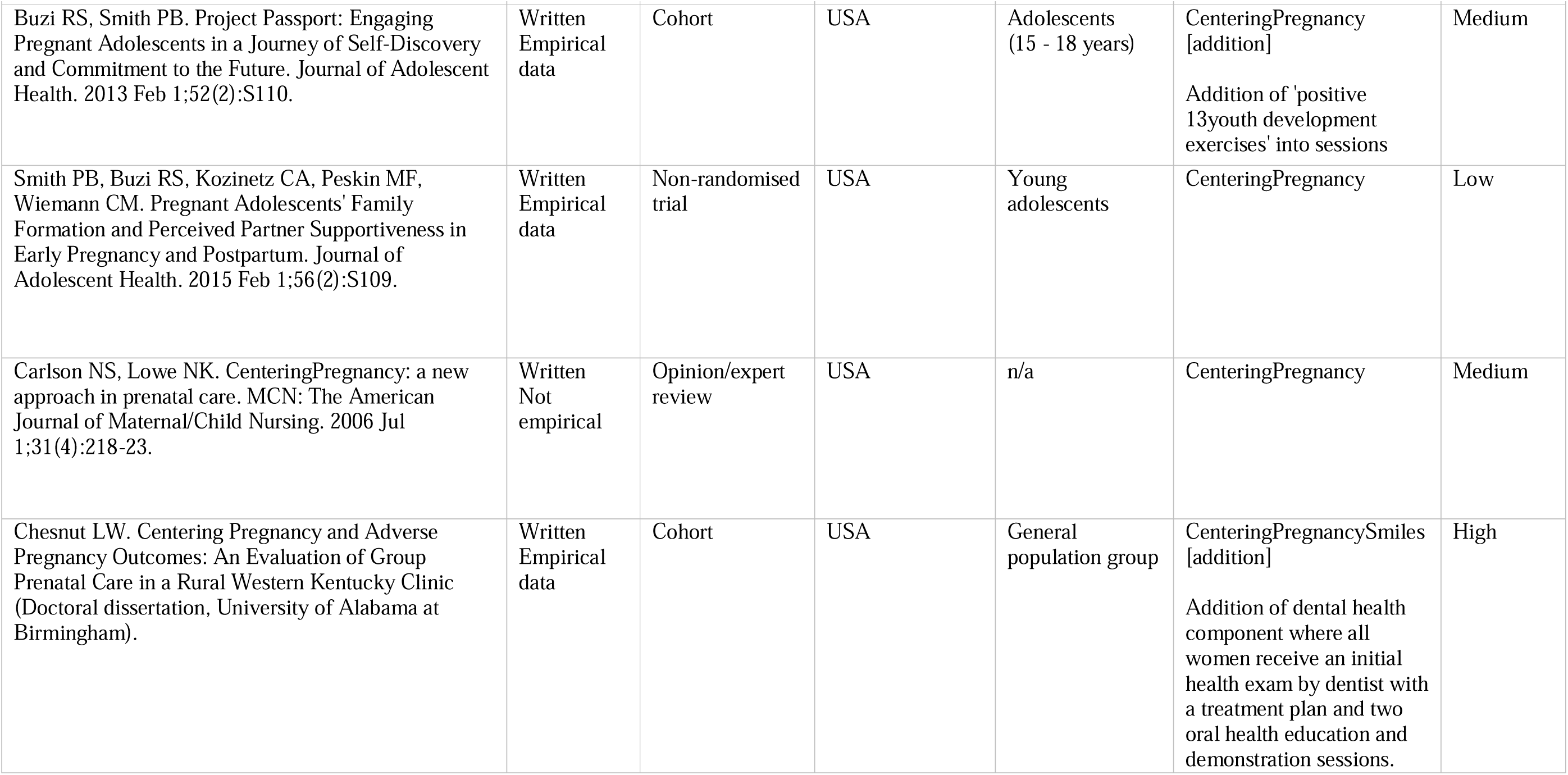
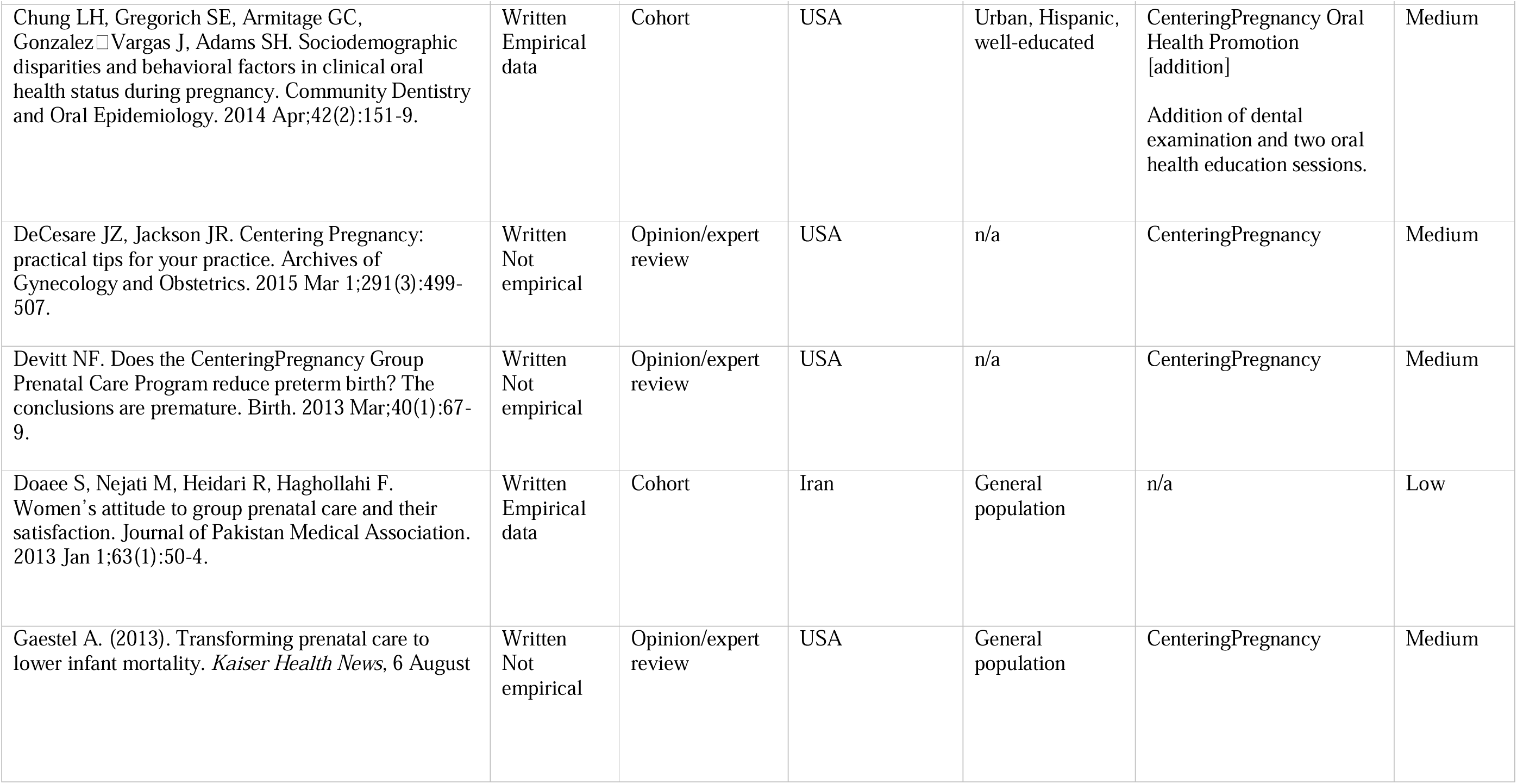
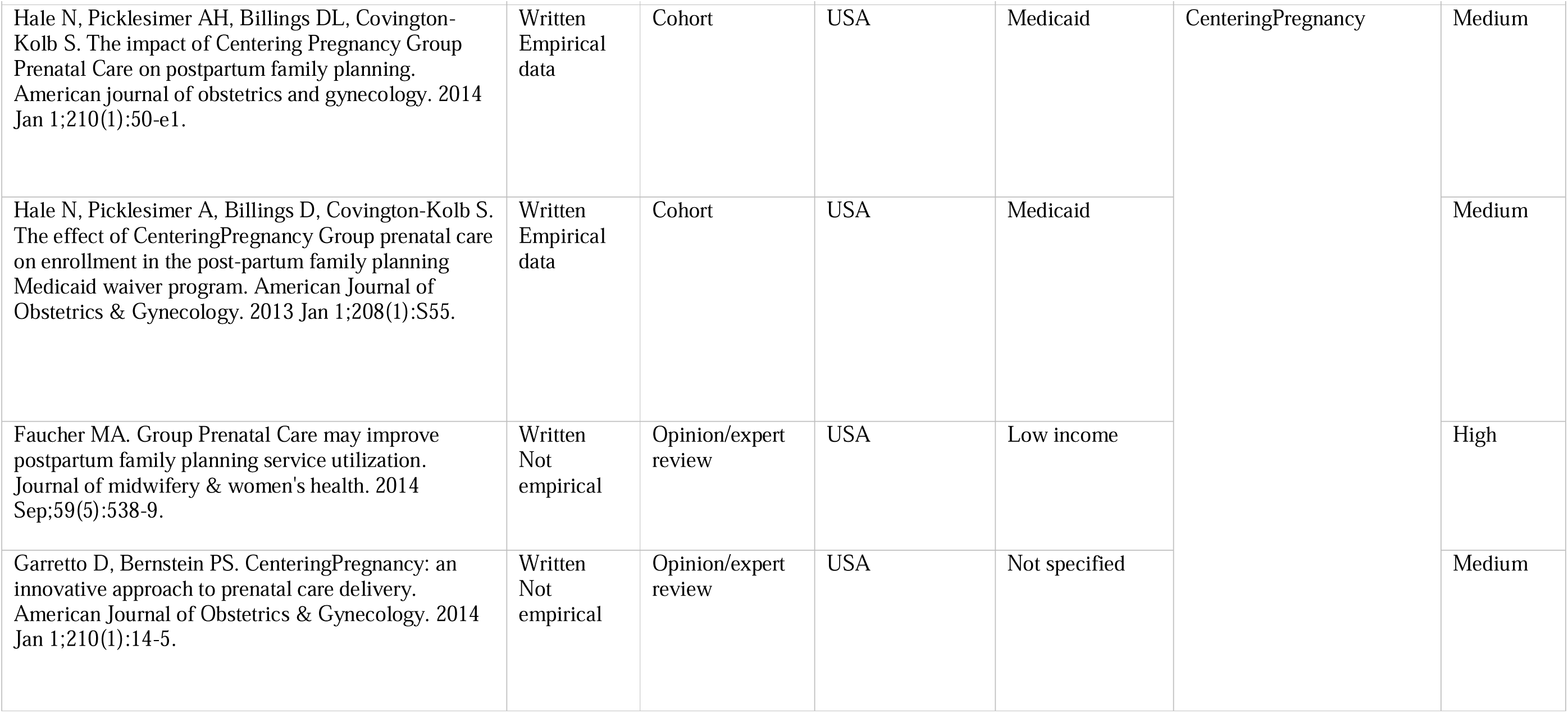
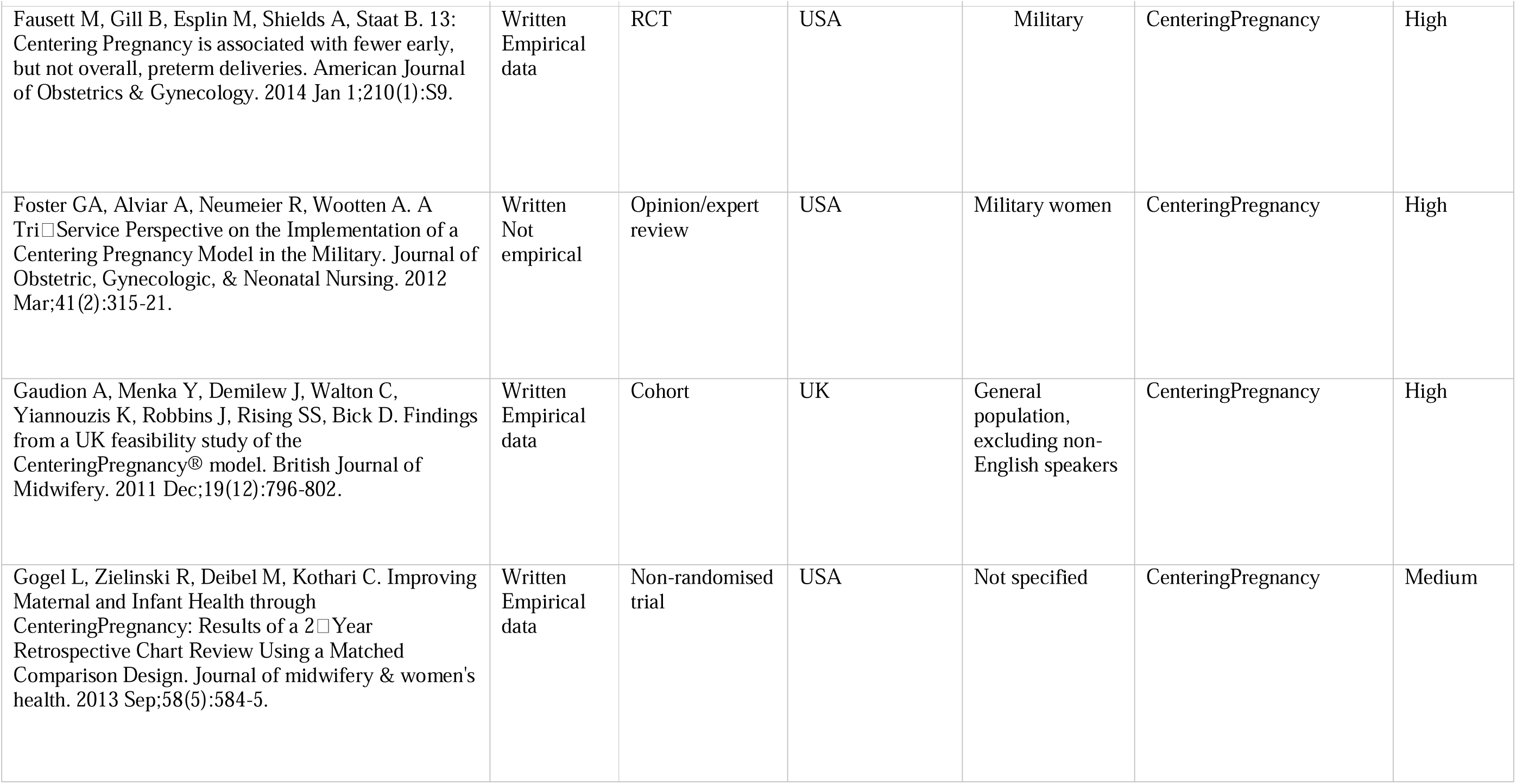
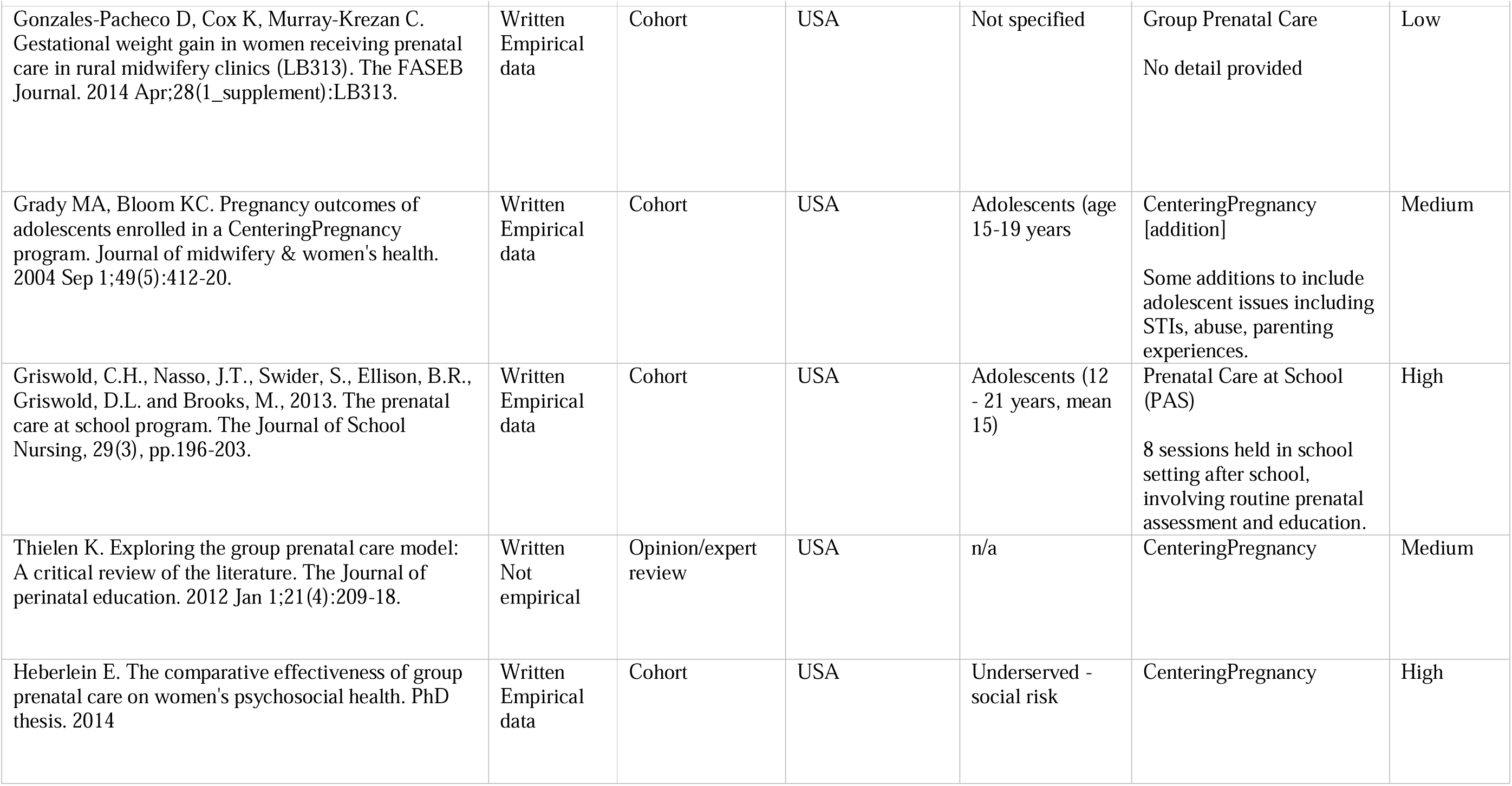
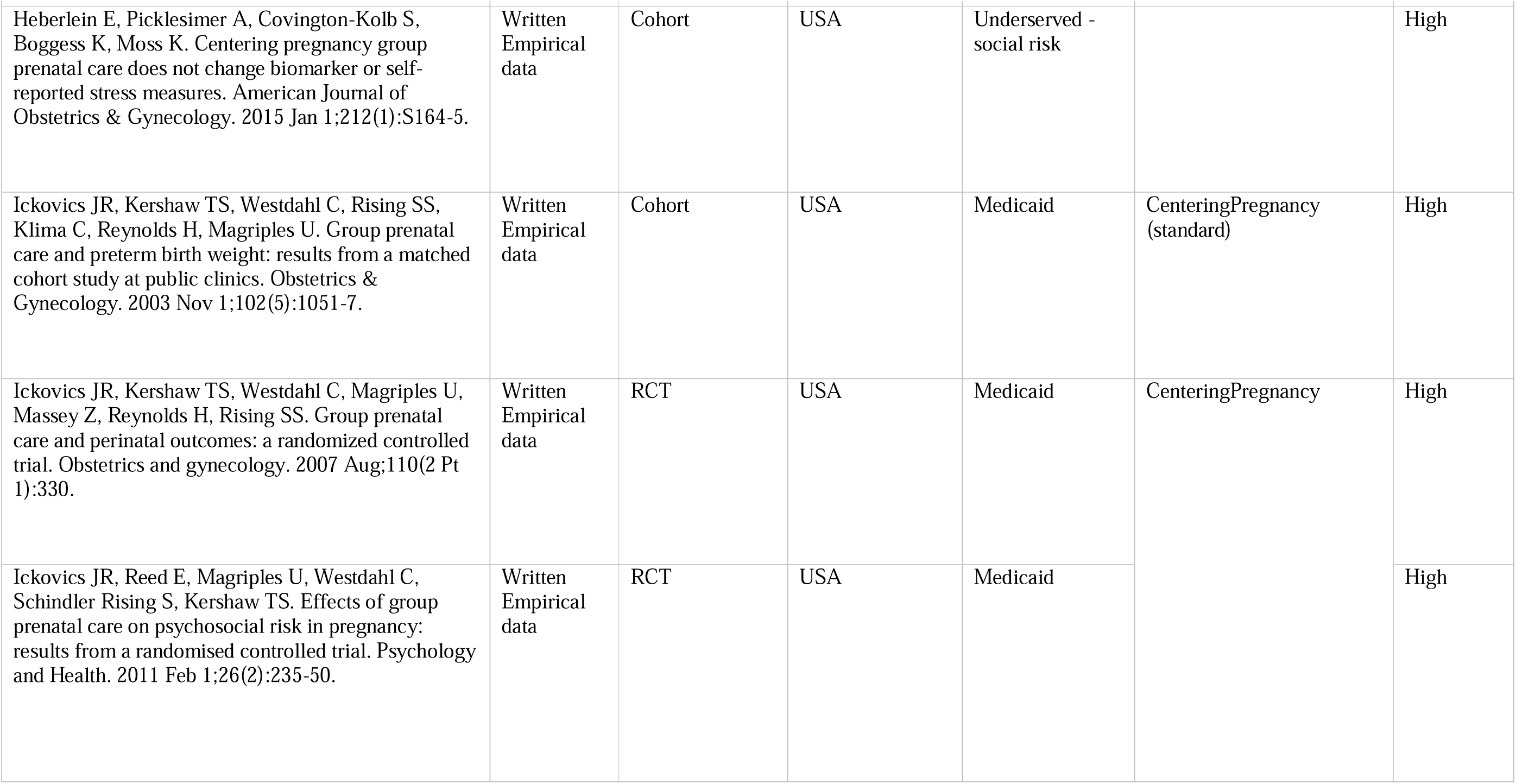
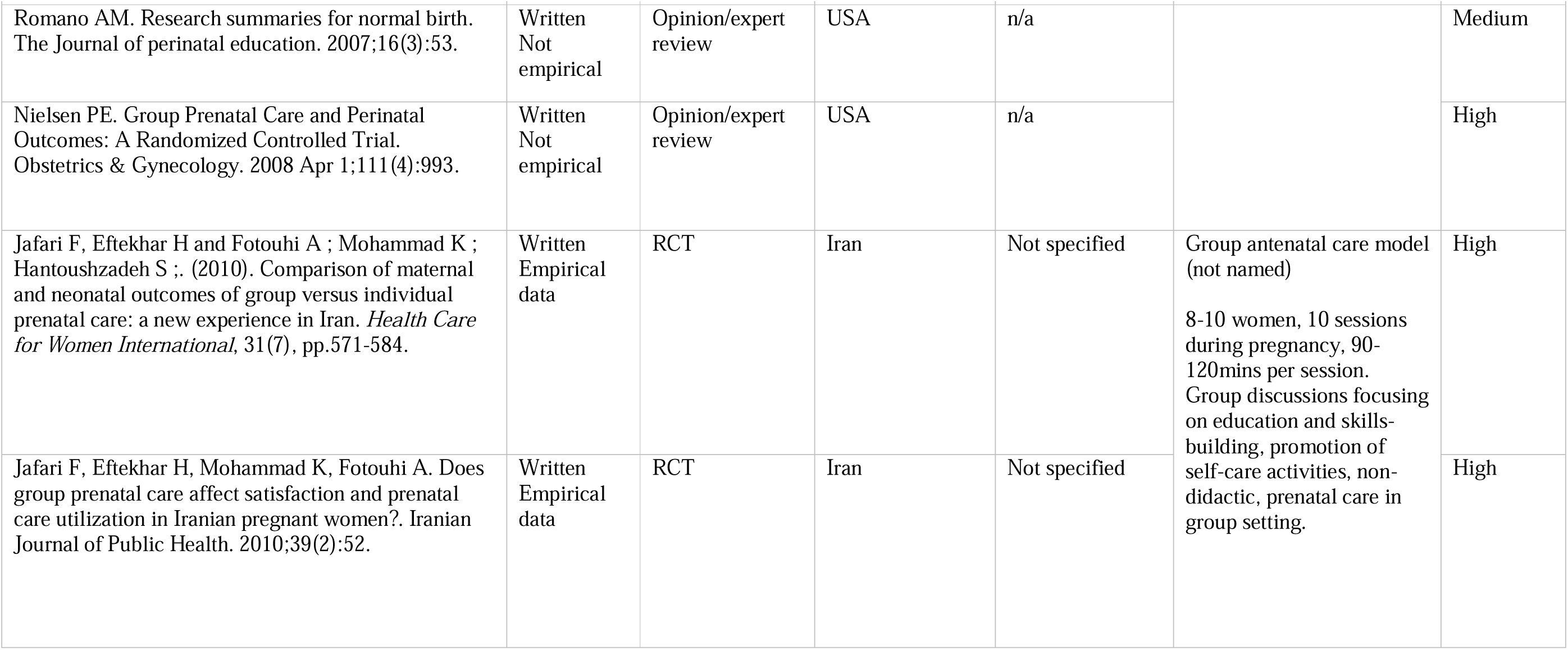
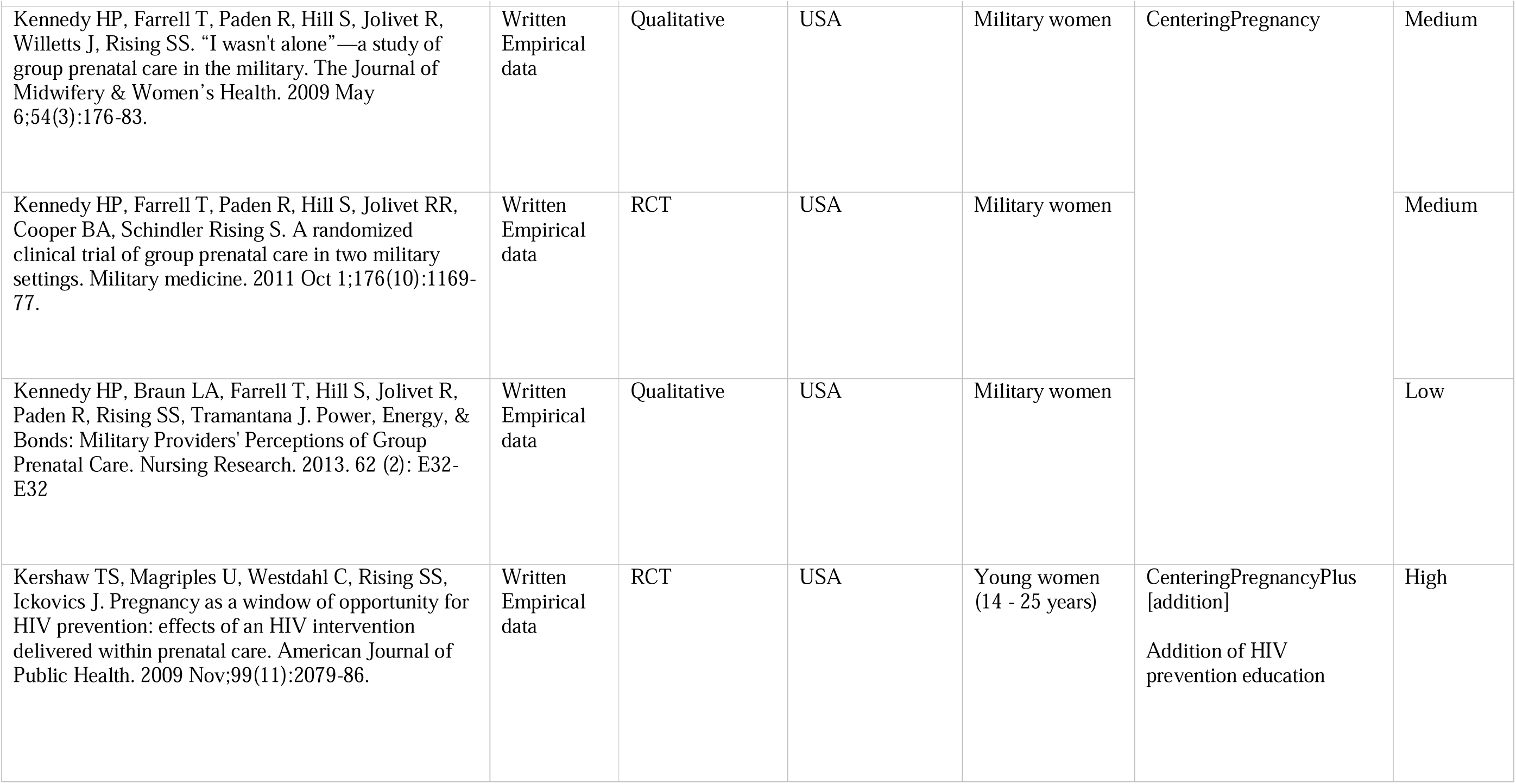
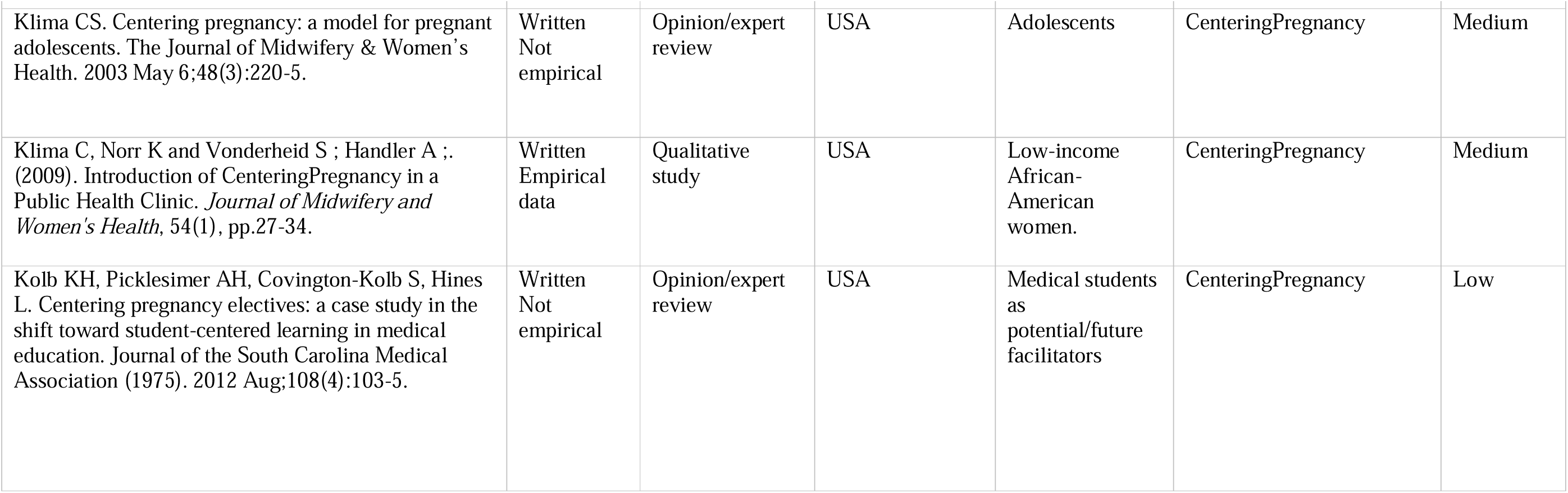
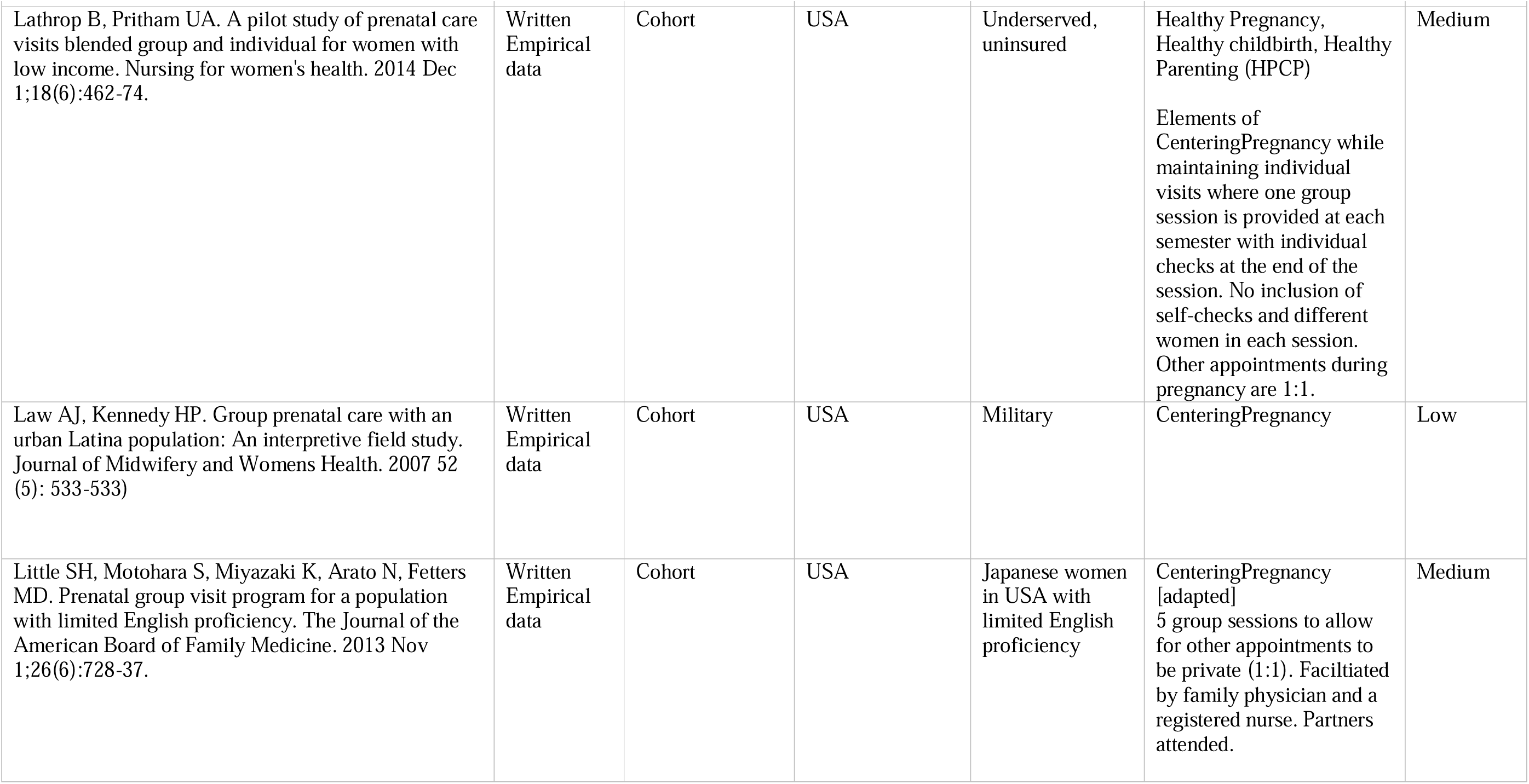
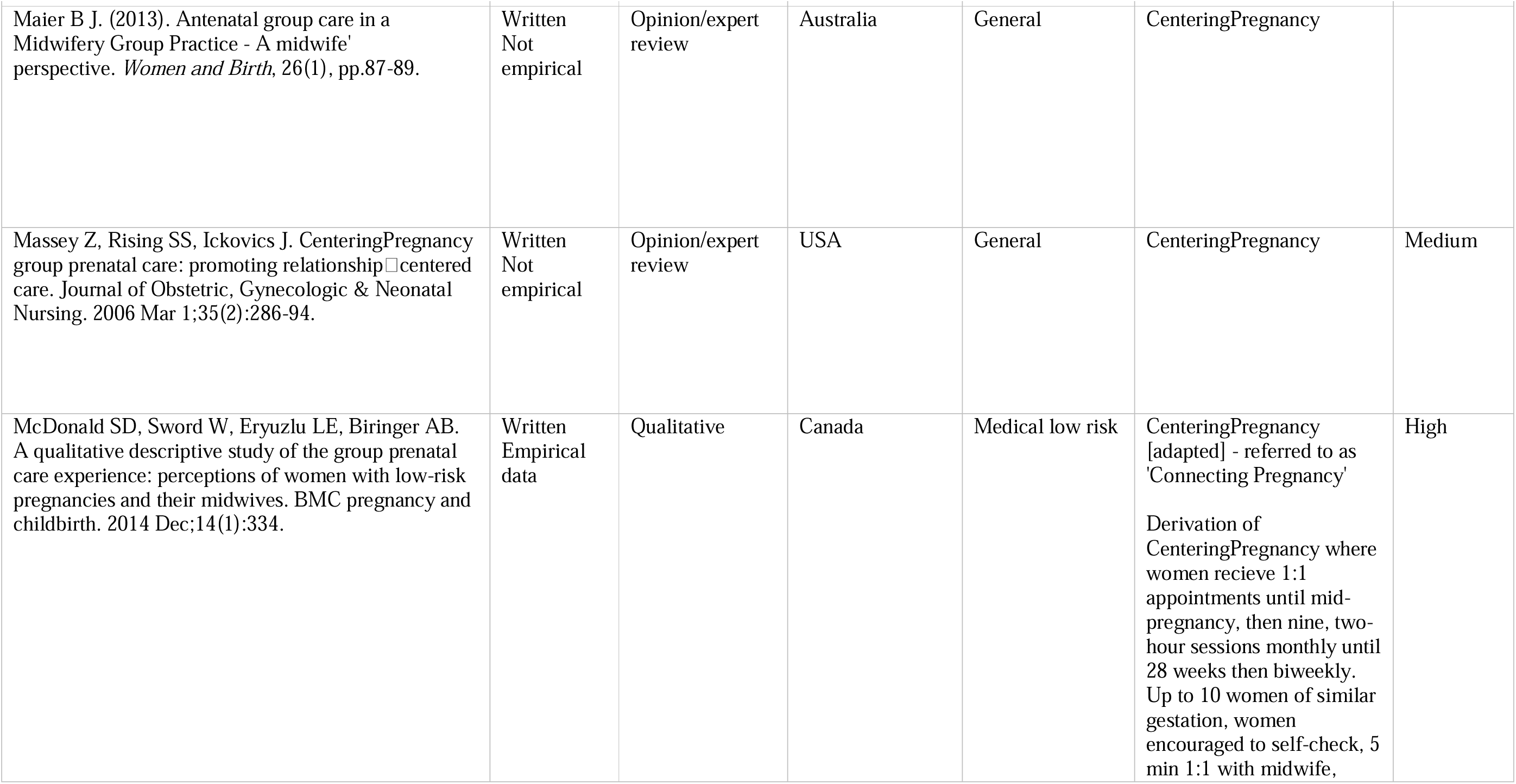
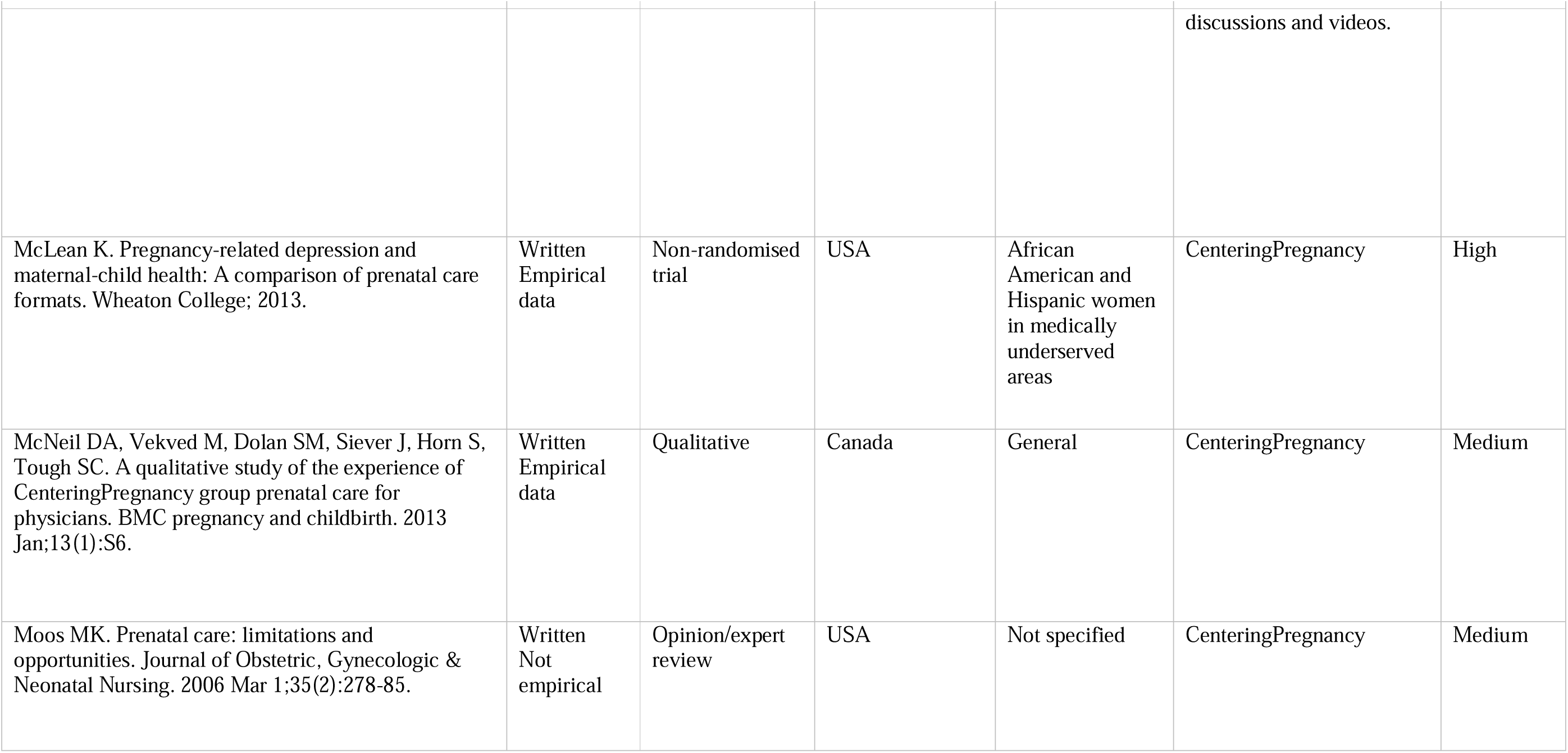
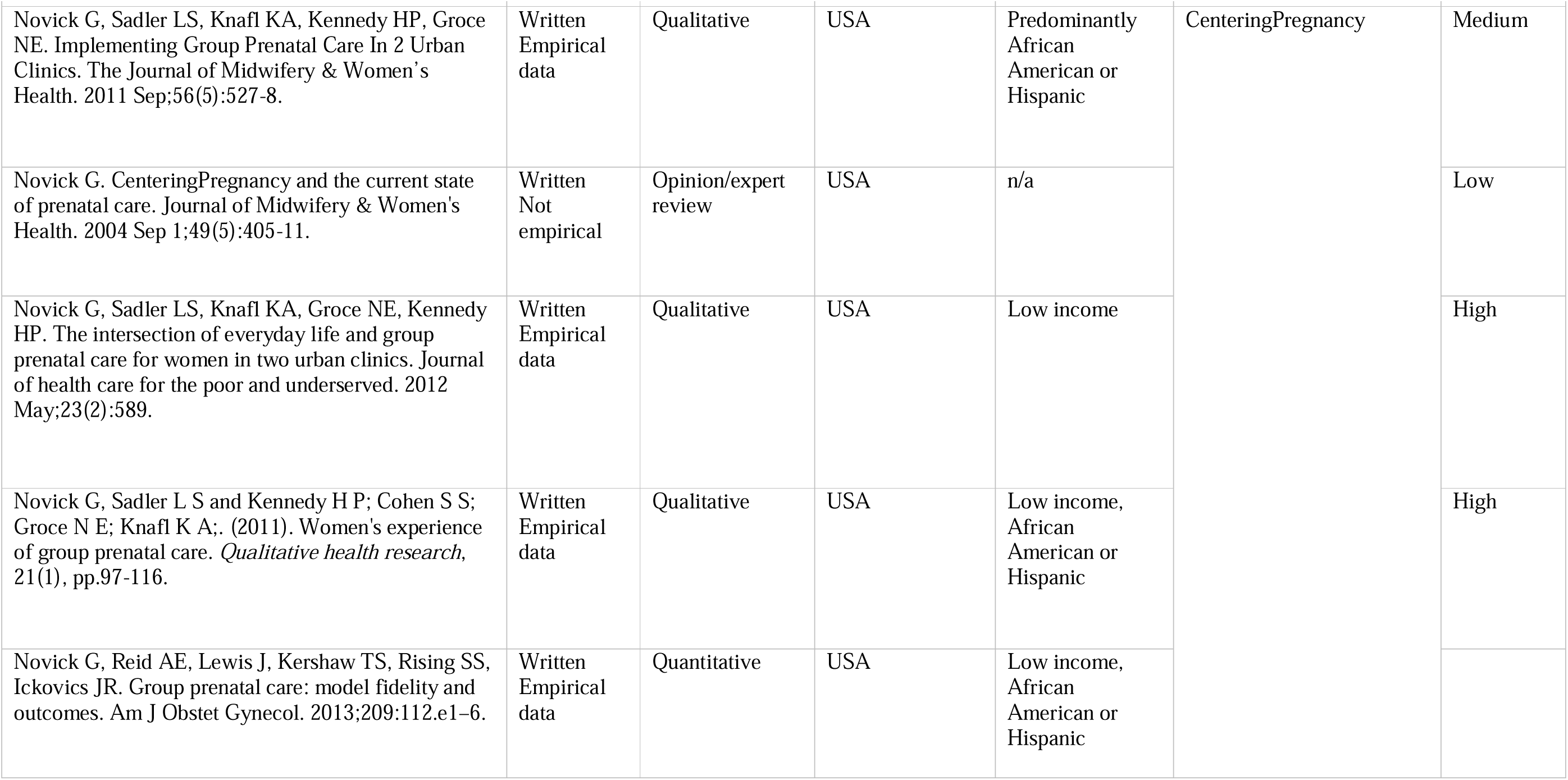
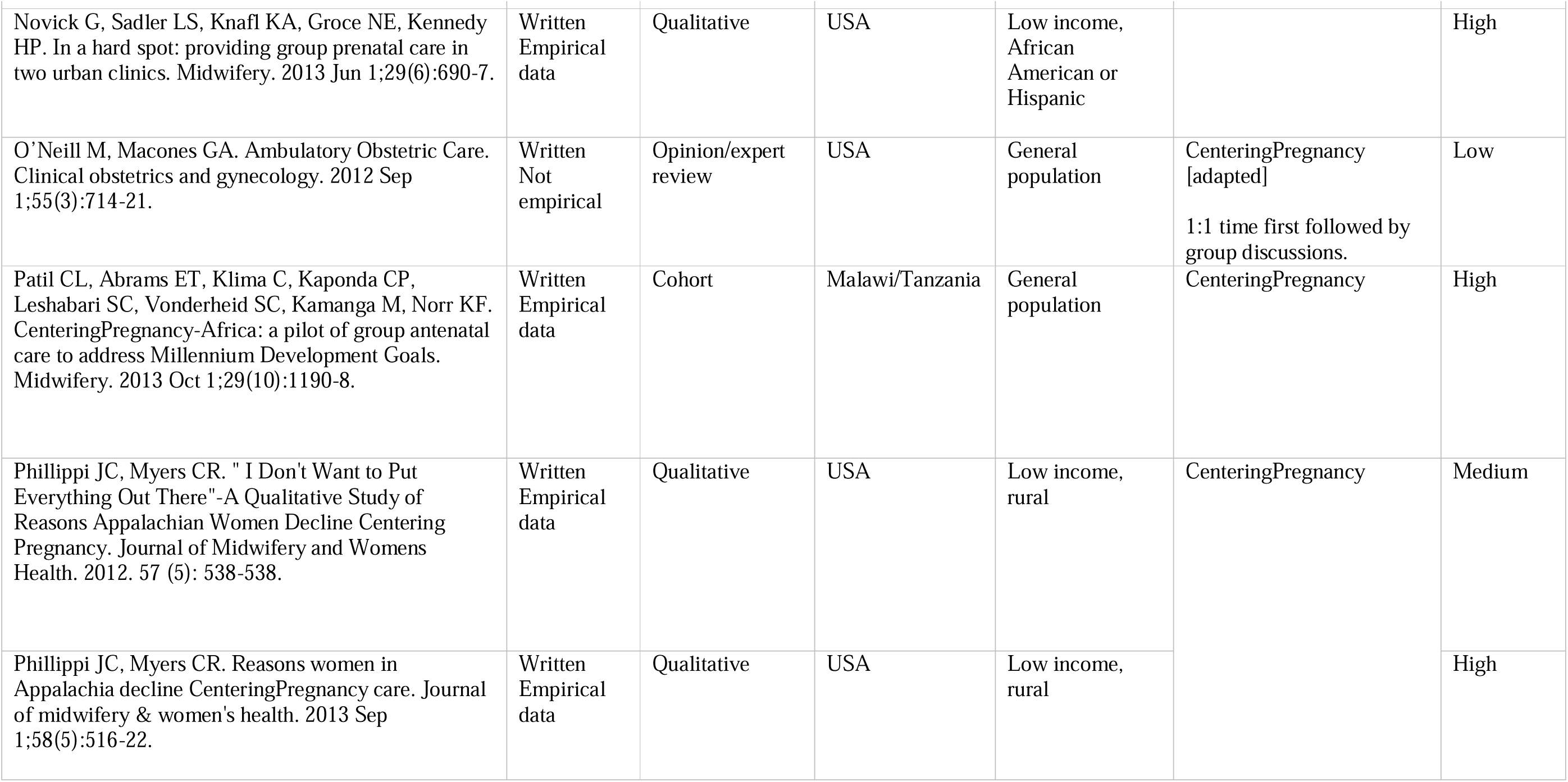
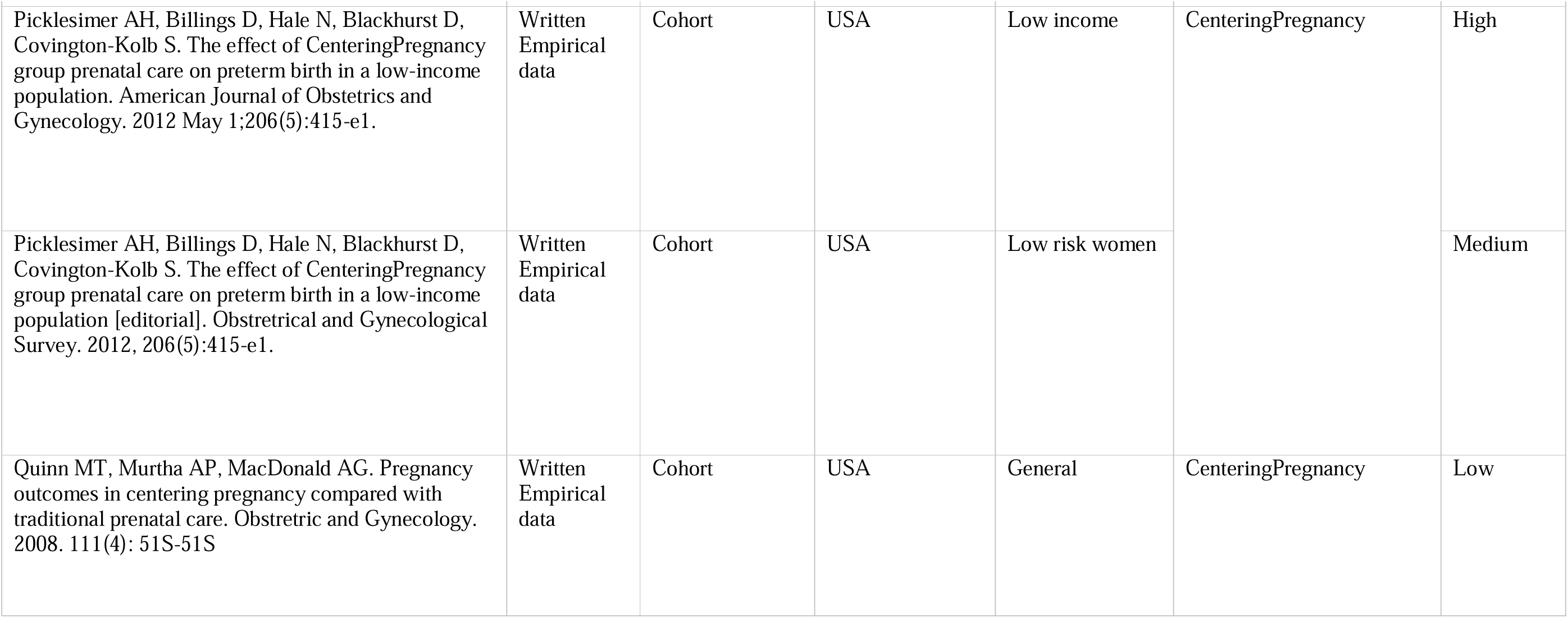
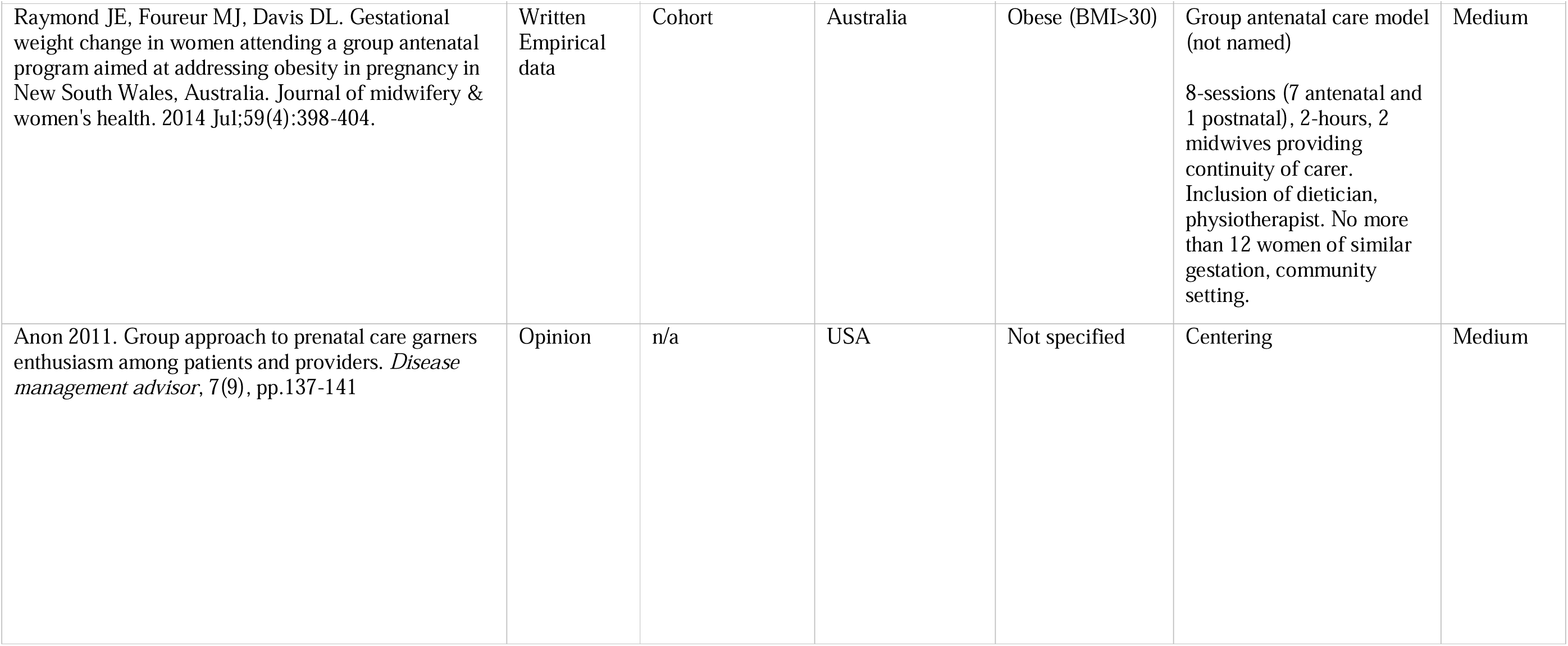
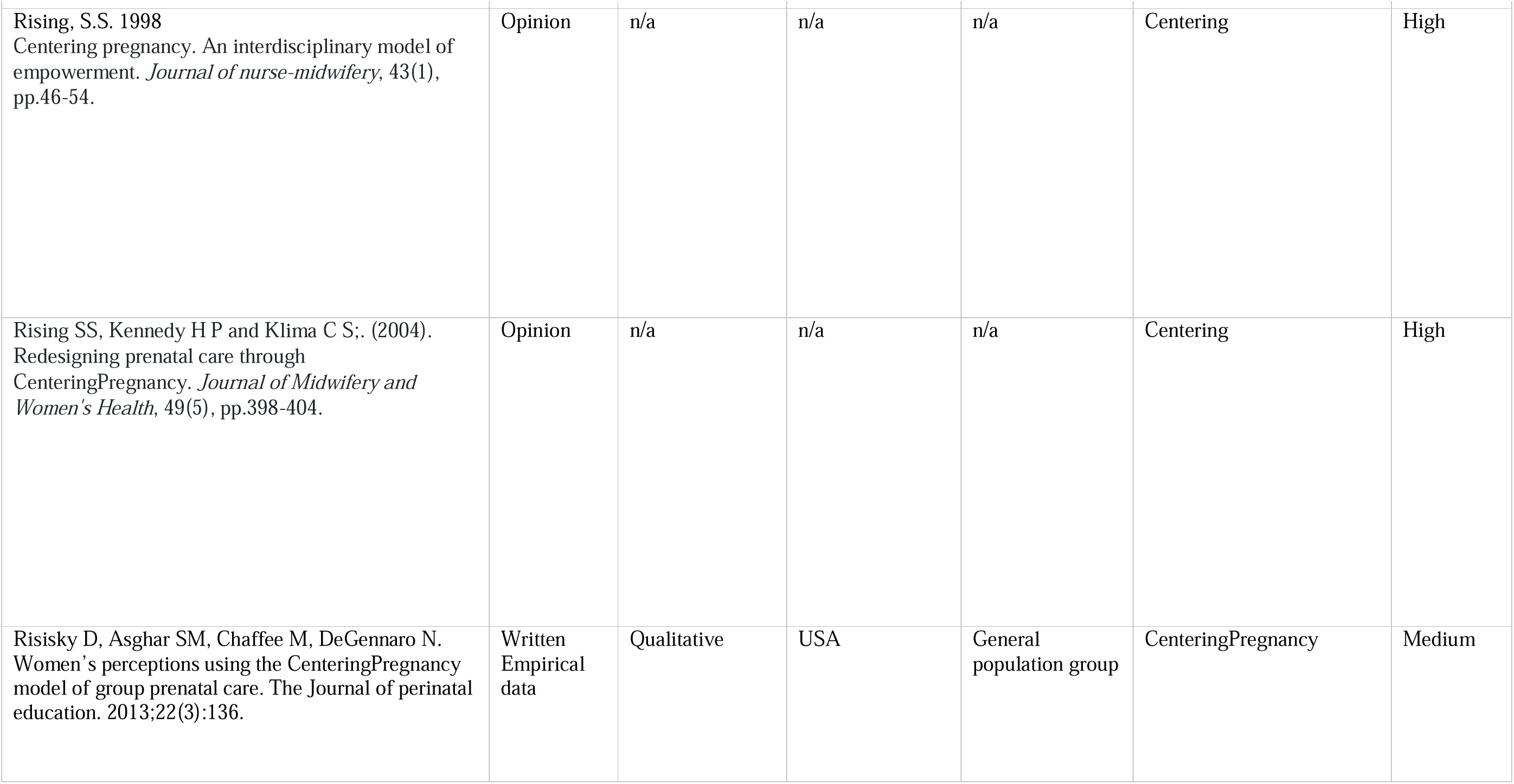
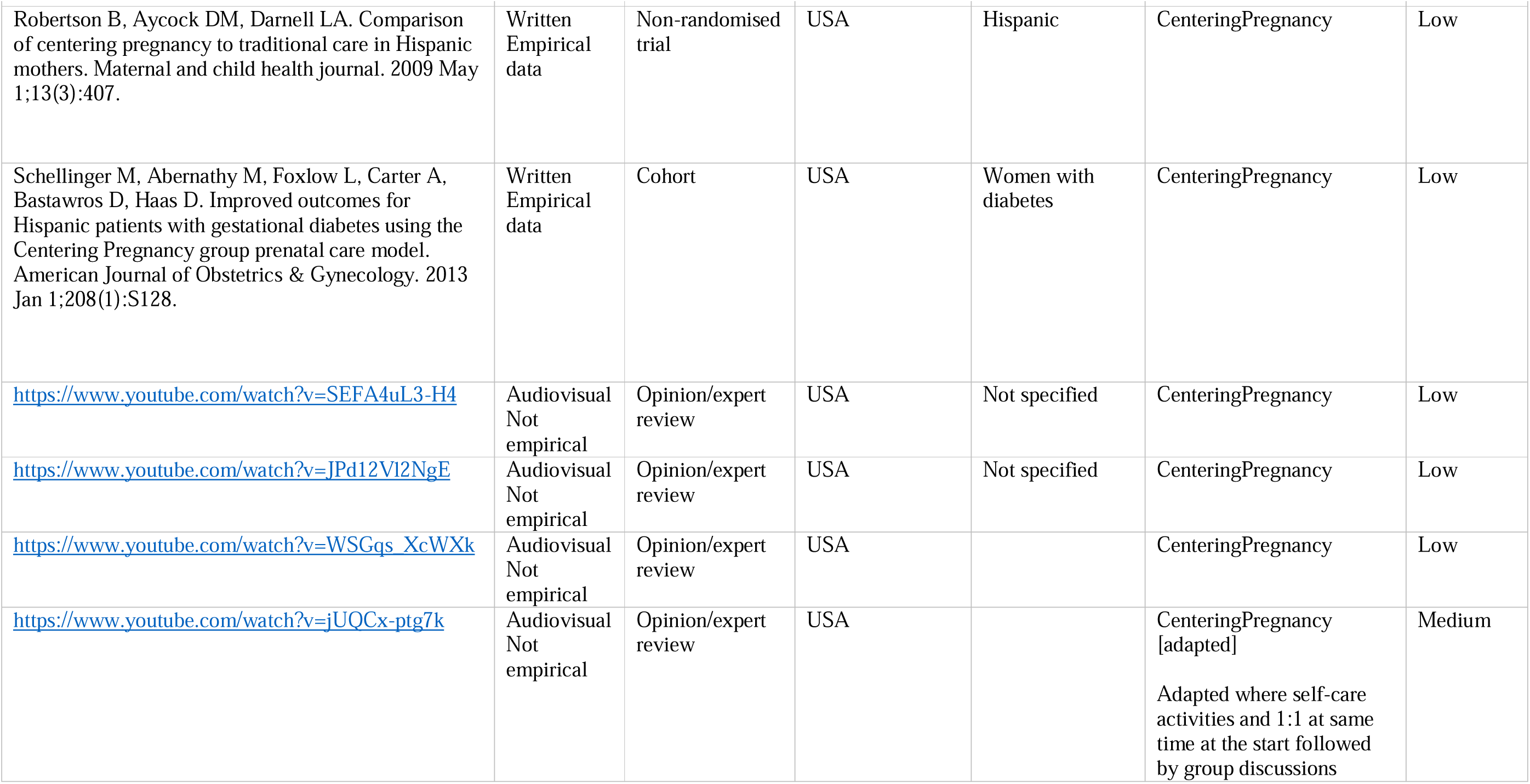
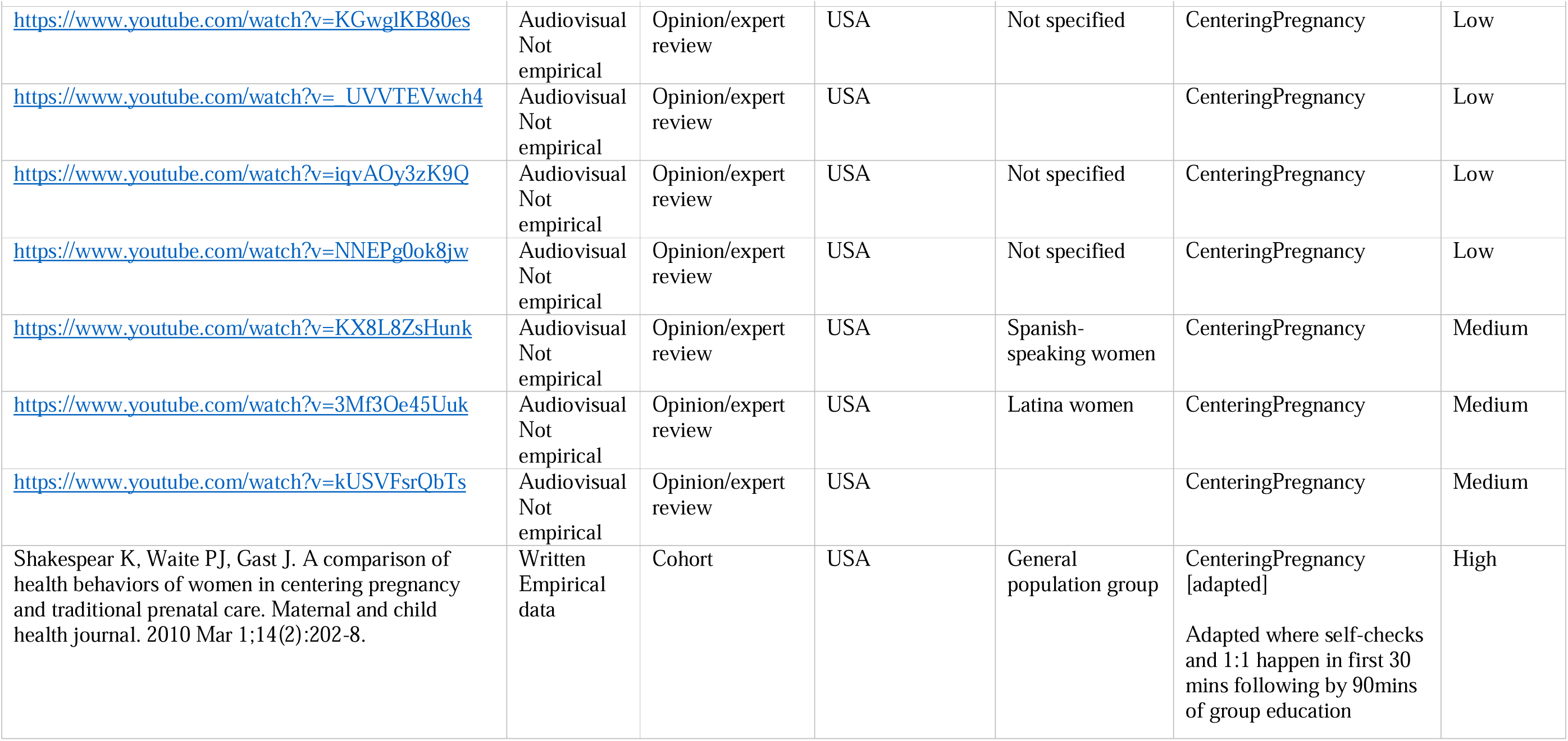
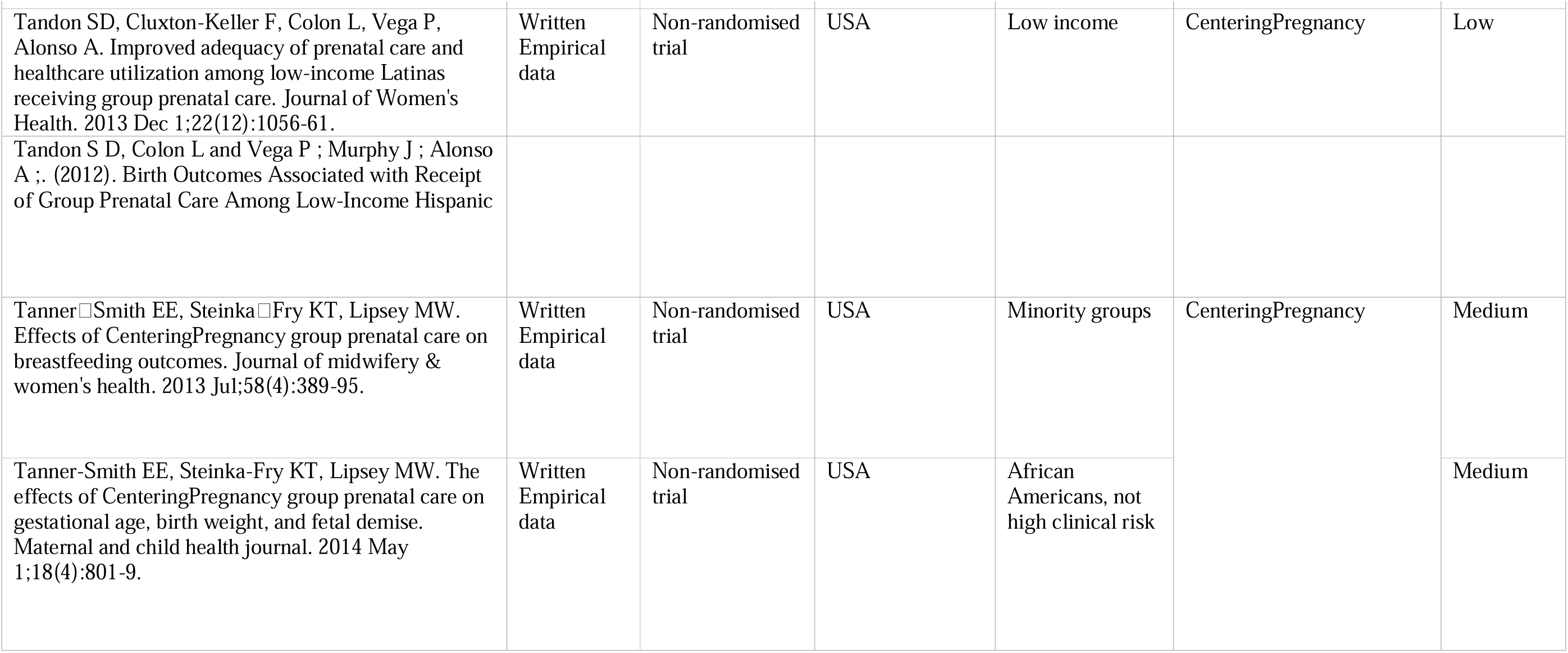
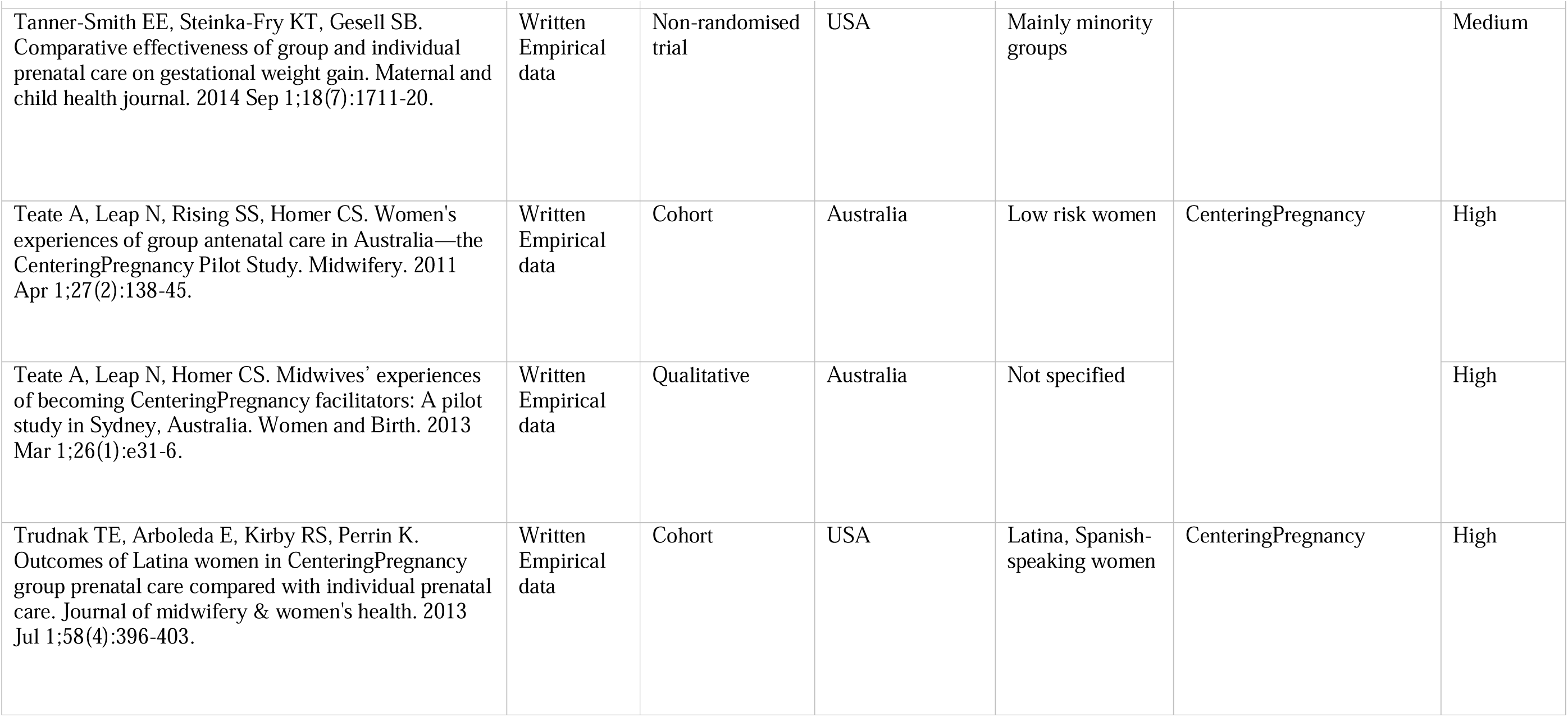
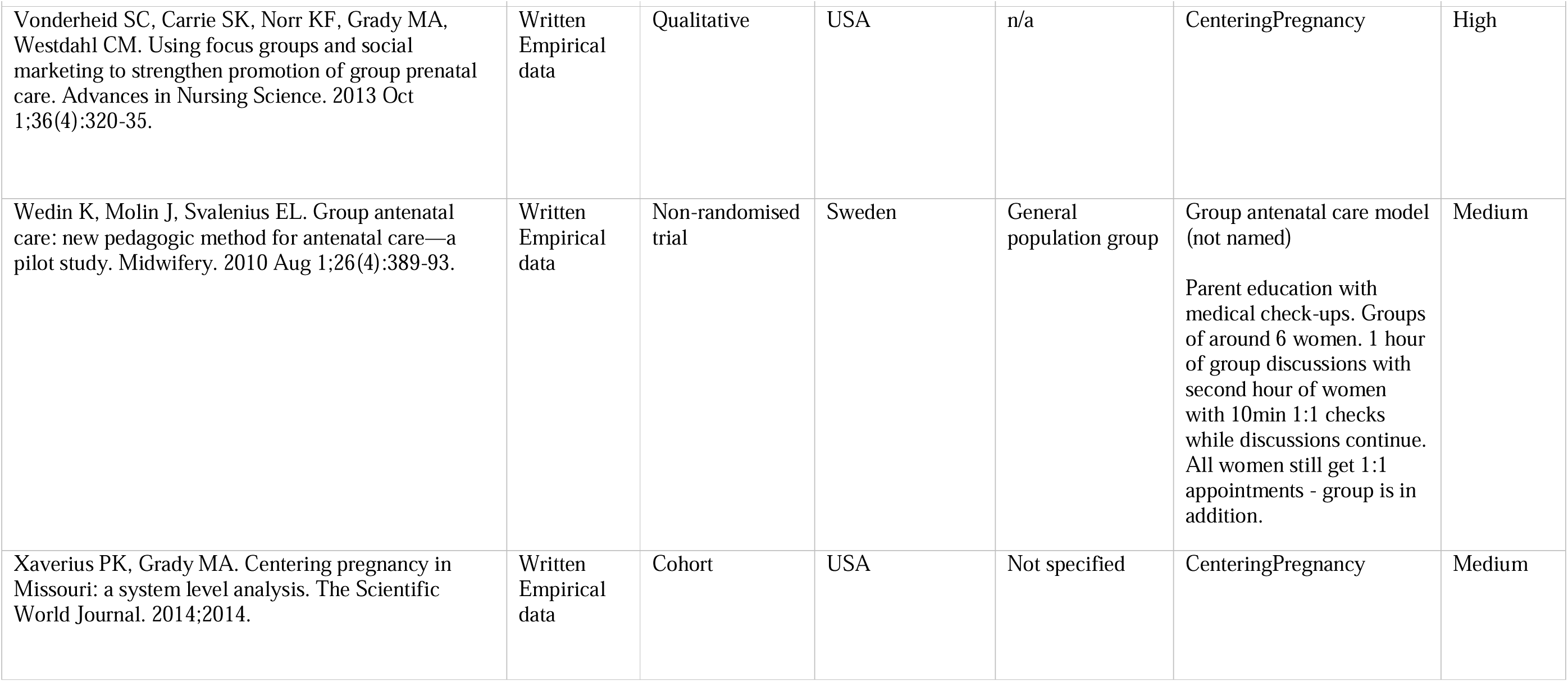
Summary table of sources included in the review.

**Supplementary file 2:** Summary table of sources included in the review update.

| Full citation | Country | Outline of study and whether any new data relevant to the review – that would amend or enhance the analysis |
| --- | --- | --- |
| Abrams, J. A., Forte, J., Bettler, C., & Maxwell, M. (2018). Considerations for implementing group-level prenatal health interventions in low-resource communities: lessons learned from Haiti. *Journal of Midwifery & Women's Health*, 63(1), 121-126. | Haiti | No detailed description of the model and how it is intended to work beyond a brief reference to findings of other studies. The main focus is how to implement this in a resource-constrained context. Challenges noted include language, literacy, space, cultural appropriateness of intervention content, and socio-political climate. |
| Adams, C., & Thomas, S. P. (2018). Alternative prenatal care interventions to alleviate Black–White maternal/infant health disparities. *Sociology compass*, 12(1), e12549. | USA | Discusses the maternal health literature that critiques standard prenatal care in the United States by drawing on intersectionality, medicalization, and fundamental causation theories. It refers to existing theories in relation to group care, including empowerment, social support and the learning from peers. The theories cited are those already identified in the review but this article's focus highlights the salience of these for black women in the US and posits that group approaches may have a particular appeal for them. |
| Brumley, J., Cain, M. A., Stern, M., & Louis, J. M. (2016). Gestational weight gain and breastfeeding outcomes in group prenatal care. *Journal of Midwifery & Women's Health*, 61(5), 557-562. | USA | A small case control study to compare gestational weight gain in women choosing and attending group prenatal care compared with standard individual prenatal care. It refers to existing theories already identified in the review including empowerment, through encouraging engagement and participation in care, and selfcare which may encourage healthy behaviours. No differences in weight gain were found but choosing group care was associated with a higher exclusive breastfeeding rate. |
| Carter, E. B., Barbier, K., Sarabia, R., Macones, G. A., Cahill, A. G., & Tuuli, M. G. (2017). Group versus traditional prenatal care in low-risk women delivering at term: a retrospective cohort study. *Journal of Perinatology*, 37(7), 769-771. | USA | Retrospective cohort study with matched controls of women with singleton, term pregnancies who participated in group care compared with traditional care. Group care was associated with a lower risk of low birth weight, of CS birth, low Apgar score or admission to higher-level neonatal care. Article included no discussion of theories relating to the hypothesised benefits, although these were covered in a 2016 systematic review by the same team. |
| Chae, S. Y., Chae, M. H., Kandula, S., & Winter, R. O. (2017). Promoting improved social support and quality of life with the CenteringPregnancy® group model of prenatal care. *Archives of women's mental health*, 20, 209-220. | USA | Prospective cohort study to assess impact of Centering on perceived social support and quality of life, finding improved scores in both. Authors refer to existing theories that underpin the hypothesised effect including social support, self-efficacy, empowerment and health education and point to the theories of social support as a stress-buffer that may reduce distress and enhance quality of life. |
| Chen, L., Crockett, A. H., Covington-Kolb, S., Heberlein, E., Zhang, L., & Sun, X. (2017). Centering and Racial Disparities (CRADLE study): rationale and design of a randomized controlled trial of centeringpregnancy and birth outcomes. *BMC pregnancy and childbirth*, 17, 1-13. | USA | Protocol for a trial of Centering pregnancy to examine impact on preterm birth. The model is hypothesised to reduce PTB and racial disparities through increased patient-provider interaction time, developing trust and understanding and empowering and promoting self-care and positive behavioural changes. |
| Cunningham SD, Lewis JB, Thomas JL, Grilo SA, Ickovics JR. Expect With Me: development and evaluation design for an innovative model of group prenatal care to improve perinatal outcomes. BMC pregnancy and childbirth. 2017 Dec;17:1-3. | USA | Account of a Group care approach with a novel integrated IT platform (which appears to be an electronic patient-held record). Authors refer to existing theories of benefit as: improved learning and skills development, attitude change and motivation, enhanced insight through sharing of common experiences and social support and facilitating development of new community norms for health-enhancing behaviours. They posit that the group approach enhances health behaviours and decision making, and helps to connect providers and patients. |
| Cunningham, S. D., Lewis, J. B., Shebl, F. M., Boyd, L. M., Robinson, M. A., Grilo, S. A., ... & Ickovics, J. R. (2019). Group prenatal care reduces risk of preterm birth and low birth weight: a matched cohort study. *Journal of women's health*, 28(1), 17-22. | USA | Retrospective matched cohort study examined the impact of group prenatal care on preterm birth and low birth weight in a large metropolitan hospital, accounting for patient adherence, over an 8.5-year period.<br>Focused on outcomes only with no additional data on theories of effect. |
| DeCesare JZ, Hannah D, Amin R. Postpartum Contraception Use Rates of Patients Participating in the Centering Pregnancy Model of Care Versus Traditional Obstetrical Care. The Journal of reproductive medicine. 2017 Jan 1;62(1-2):45-9. | USA | Retrospective chart review focused on postpartum contraceptive rates<br>Focused on outcomes only.<br>No relevant data included. |
| Earnshaw VA, Rosenthal L, Cunningham SD, Kershaw T, Lewis J, Rising SS, Stasko E, Tobin J, Ickovics JR. Exploring group composition among young, urban women of color in prenatal care: Implications for satisfaction, engagement, and group attendance. *Women's Health Issues*. 2016 Jan 1;26(1):110-5. | USA | Analyses explored composition of groups in terms of age, race, ethnicity, and language, suggesting diverse age composition seems to be associated with young women's engagement in care, including young women of colour.<br>No new theories but useful exploration regarding group composition. |
| Eluwa GI, Adebajo SB, Torpey K, Shittu O, Abdu-Aguye S, Pearlman D, Bawa U, Olorukooba A, Khamofu H, Chiegli R. The effects of centering pregnancy on maternal and fetal outcomes in northern Nigeria; a prospective cohort analysis. BMC Pregnancy and Childbirth. 2018 Dec;18(1):1-0. | Nigeria | Prospective cohort study assessed the effect of Centering pregnancy groups on the uptake of antenatal care, facility delivery and immunization rates for infants.<br>No relevant theory data included. |
| Felder JN, Epel E, Lewis JB, Cunningham SD, Tobin JN, Rising SS, Thomas M, Ickovics JR. Depressive symptoms and gestational length among pregnant adolescents: Cluster randomized control trial of CenteringPregnancy® plus group prenatal care. *Journal of consulting and clinical psychology*. 2017 Jun;85(6):574. | USA | Cluster randomised trial focused on whether this model can reduce depressive symptoms in adolescents, given the clinical benefits found for adolescent mothers in some studies. Refers to existing evidence that group care provides opportunities for social support, and that participants learn stress reduction techniques and communication skills, which may improve psychosocial functioning. Significant reductions in depressive symptoms were observed and these may be associated with rates of preterm birth and gestational age at birth. |
| Fuentes-Rivera E, Heredia-Pi I, Andrade-Romo Z, Alcalde-Rabanal J, Bravo L, Jurkiewicz L, Darney BG. Evaluating process fidelity during the implementation of Group Antenatal Care in Mexico. BMC health services research. 2020 Dec;20:1-8. | Mexico | Descriptive study of model fidelity after implementation. Refers to existing theories and examines fidelity to published features of the Centering model, using a checklist. Overall fidelity was high but with some variability.<br>No relevant theory data included. |
| Gareau S, Lopez-De Fede A, Loudermilk BL, Cummings TH, Hardin JW, Picklesimer AH, Crouch E, Covington-Kolb S. Group prenatal care results in Medicaid savings with better outcomes: a propensity score analysis of CenteringPregnancy participation in South Carolina. Maternal and child health journal. 2016 Jul;20:1384-93. | USA | Retrospective cohort study of Centering Pregnancy in Medicare in US found cost savings relating to prevention of adverse birth outcomes.<br>No relevant theory data included. |
| Gennaro S, Melnyk BM, O'Connor C, Gibeau AM, Nadel E. Improving prenatal care for minority women. MCN. The American journal of maternal child nursing. 2016 May;41(3):147. | USA | Discussion article on range of models of care which notes that group care may address health behaviours relating to physical and mental health, which may 'decrease some of the disparities in birth outcomes that are well documented between minority and majority women, as minority women are known to experience increased levels of stress, anxiety, and depression.' |
| Gholipour K, Tabrizi JS, Asghari Jafarabadi M, Iezadi S, Mardi A. Effects of customer self-audit on the quality of maternity care in Tabriz: a cluster-randomized controlled trial. PLoS One. 2018 Oct 11;13(10):e0203255. | Iran | Customer self-audit of CenteringPregnancy® or usual care within the context of a RCT. The intervention group scored higher on the Service Quality aspects confidentiality, communication, autonomy, availability of support group, dignity, safety, prevention, and accessibility.<br>No account of the model itself apart from referring to existing evidence on satisfaction and potential empowerment. Findings suggest it may enhance empowerment for this clientele. |
| Grant JH, Handwerk K, Baker K, Milling V, Barlow S, Vladutiu CJ. Implementing Group Prenatal Care in Southwest Georgia Through Public–Private Partnerships. Maternal and Child Health Journal. 2018 Nov;22:1535-42. | USA | Describes the successful implementation of CenteringPregnancy in a public health setting with no prior prenatal services; assesses the program's first 5-year perinatal outcomes; and discusses several key lessons learned using review of medical records. Refers to previous work showing improved satisfaction and uptake of care but no further information on model. Reports success in implementing and increasing attendance rates. |
| Heberlein EC, Frongillo EA, Picklesimer AH, Covington-Kolb S. Effects of group prenatal care on food insecurity during late pregnancy and early postpartum. Maternal and child health journal. 2016 May;20:1014-24.<br>Heberlein EC, Picklesimer AH, Billings DL, Covington□ Kolb S, Farber N, Frongillo EA. Qualitative comparison of women's perspectives on the functions and benefits of group and individual | USA | Further papers from a doctoral study included in the original review: full data of theories obtained from PhD thesis in original analysis. The additional papers corroborate earlier reports of positive impact on stress, confidence, knowledge, motivation, informed decision making, and health care engagement rather than adding new theoretical information or findings |
| prenatal care. Journal of midwifery & women's health. 2016 Mar;61(2):224-34.<br>Heberlein EC, Picklesimer AH, Billings DL, Covington-Kolb S, Farber N, Frongillo EA. The comparative effects of group prenatal care on psychosocial outcomes. Archives of women's mental health. 2016 Apr;19:259-69. |  |  |
| Heredia□Pi IB, Fuentes□Rivera E, Andrade□Romo Z, Bravo Bolaños Cacho MD, Alcalde□Rabanal J, Jurkiewicz L, Darney BG. The Mexican experience adapting CenteringPregnancy: lessons learned in a publicly funded health care system serving vulnerable women. Journal of midwifery & women's health. 2018 Sep;63(5):602-10. | Mexico | Implementation focused account of adapting the standard Centering Model for the Mexican context. Basic account of Centering and refers to the challenge of moving towards a more facilitative model in a hierarchical medical context. |
| Hetherington E, Tough S, McNeil D, Bayrampour H, Metcalfe A. Vulnerable women's perceptions of individual versus group prenatal care: results of a cross-sectional survey. Maternal and Child Health Journal. 2018 Nov;22:1632-8. | Canada | This survey aimed to assess patient experience among vulnerable women in group prenatal care compared to individual care. Reports that women felt better informed and that providers were more interested in them. |
| Hodgson ZG, Saxell L, Christians JK. An evaluation of Interprofessional group antenatal care: a prospective comparative study. BMC Pregnancy and childbirth. 2017 Dec;17:1-9. | Canada | Cohort study and survey compares outcomes in women receiving interprofessional group perinatal care 'Connecting Pregnancy' versus interprofessional individual care (nurses, midwives and physicians, plus sometimes guests such as doulas). Group care considered similar in this programme but with potential to enhance quality of information. |
| Ickovics JR, Earnshaw V, Lewis JB, Kershaw TS, Magriples U, Stasko E, Rising SS, Cassells A, Cunningham S, Bernstein P, Tobin JN. Cluster randomized controlled trial of group prenatal care: perinatal outcomes among adolescents in New York City health centers. American journal of public health. 2016 Feb;106(2):359-65. | USA | RCT of Perinatal outcomes. Women at intervention sites were significantly less likely to have infants small for gestational age; women with more group visits had improvements in gestational age, birth weight, days in neonatal intensive care unit, rapid repeat pregnancy, condom use, and unprotected sex with no associated risks. No additional data on the model or theory of effect but identified improvements for adolescents. |
| Jensen MN, Fage-Butler AM. Antenatal group consultations: Facilitating patient-patient education. Patient Education and Counseling. 2016 Dec 1;99(12):1999-2004. | Denmark | Qualitative study focused on understanding whether and how peer learning is facilitated in group settings; peer learning conceptualised as patient-patient education. Main focus of gANC in Denmark is described as health promotion and it is hypothesised that peer-learning enhances this, via practical, experiential and emotional knowledge. 'Groupness' of the group was considered to enhance peer learning and did not depend on homogeneity because of shared experience of |
|  |  | pregnancy/birth, along with facilitative skills of the midwife. |
| Jolivet RR, Uttekar BV, O'Connor M, Lakhwani K, Sharma J, Wegner MN. Exploring perceptions of group antenatal Care in Urban India: results of a feasibility study. Reproductive health. 2018 Dec;15(1):1-1. | India | Focused on potential implementation of a generic model of GANC in India, combined from Centering Pregnancy and the Home-based Lifesaving Skills Programme, aligned with local guidelines. No detail of description of theories. Gathered provider, women and families' views about acceptability and feasibility, based on a single demonstration in 3 health-service settings. GANC was seen as having potential to improve experiences of care, empower women to become more active partners and participants in their care, through self-assessment, active learning and peer support. |
| Kabue MM, Grenier L, Suhowatsky S, Oyetunji J, Ugwa E, Onguti B, Omanga E, Gichangi A, Wambua J, Waka C, Enne J. Group versus individual antenatal and first year postpartum care: Study protocol for a multi-country cluster randomized controlled trial in Kenya and Nigeria. Gates Open Research. 2018;2. | Kenya and Nigeria | Protocol for a RCT comparing GANC and individual care in two African countries. Refers only to general aim of improving care uptake and outcomes and to test the model in a low-income country context. |
| Kania-Richmond A, Hetherington E, McNeil D, Bayrampour H, Tough S, Metcalfe A. The impact of introducing centering pregnancy in a community health setting: a qualitative study of experiences and perspectives of health center clinical and support staff. Maternal and Child Health Journal. 2017 Jun;21:1327-35. | Canada | Focused on staff experience and perceptions of integration in an existing clinic setting, challenges and opportunities. Centering Pregnancy; No specific focus on the model itself. |
| Kearney L, Kynn M, Craswell A, Reed R. The relationship between midwife-led group-based versus conventional antenatal care and mode of birth: a matched cohort study. BMC pregnancy and childbirth. 2017 Dec;17(1):1-7. | Australia | Retrospective matched cohort study focused on clinical birth outcomes. Non-significant association with reduced rate of CS was found. Basic description of the model 'Expecting and Connecting' as interactive, less didactic and facilitating midwife continuity of carer antenatally; related theories not discussed but previous evidence of impact was reviewed briefly. |
| Kominiarek MA, Crockett A, Covington-Kolb MS, Simon M, Grobman WA. Association of group prenatal care with gestational weight gain. Obstetrics and gynecology. 2017 Apr;129(4):663. | USA | Retrospective cohort study of association of group prenatal care (Centering Pregnancy) with gestational weight gain. Hypothesises that the focus on skills, social support and self-monitoring may assist weight management. No differences were found but this could have been related to confounding factors. |
| Kweekel L, Gerrits T, Rijnders M, Brown P. The role of trust in centeringpregnancy: building interpersonal trust relationships in group-based prenatal care in The Netherlands. Birth. 2017 Mar;44(1):41-7. | The Netherlands | Qualitative study with women who had experienced Centering Pregnancy. Concept of trust was identified as a key mechanism enhancing the group processes of social support and sharing. 'Trust facilitated social support which in turn enabled reassurance and the building of women's self-confidence.'<br>Examines the mechanisms that create trusting relationships within CP to better understand CP outcomes and effectiveness. |
|  |  | Enhances existing analysis. |
| Little, S. H., & Feters, M. D. (2019). Transcultural modifications of a Japanese language group prenatal care program for transcultural adaptation. *Journal of Transcultural Nursing*, 30(2), 106-114. | Japan | Account of the transcultural adaptation of a group prenatal care program for Japanese women. Refers only to existing evidence on benefits. Adaptations were mainly in detail of discussion content apart from adaptations to increase privacy: use of a divider screen and avoidance of discussion of sensitive topics in group visits; reduction in number of group visits from 10 to 6. |
| Lori JR, Ofosu-Darkwah H, Boyd CJ, Banerjee T, Adanu RM. Improving health literacy through group antenatal care: a prospective cohort study. *BMC Pregnancy and Childbirth*. 2017 Dec;17(1):1-9. | Ghana | Cohort study to assess impact of group care on health literacy, conceptualised as ability to understand and act on health messages from professionals, especially relating to risk warning signs and breastfeeding. Time and an interactive and patient-centred approach to information giving were hypothesised as contributors to enhanced literacy. |
| Lori JR, Chuey M, Munro-Kramer ML, Ofosu-Darkwah H, Adanu RM. Increasing postpartum family planning uptake through group antenatal care: a longitudinal prospective cohort design. *Reproductive health*. 2018 Dec;15:1-8. | Ghana | Prospective cohort study. Women were followed for one-year postpartum to examine the uptake and continuation of family planning following enrolment in group versus individual ANC. Family Planning uptake and breastfeeding rates found to be improved. Hypothesis based on view that in GANC women would have more time to gain and process information about postnatal health issues. |
| Liu R, Chao MT, Jostad-Laswell A, Duncan LG. Does CenteringPregnancy group prenatal care affect the birth experience of underserved women? A mixed methods analysis. *Journal of immigrant and minority health*. 2017 Apr;19:415-22. | USA | Qualitative study focused on birth experiences of immigrant and minority women and how CenteringPregnancy may have influenced these. Model is examined through reference to existing CP literature although they highlight the concept of self-efficacy as being central. A mindfulness approach was incorporated but this is not explained. Centering was reported as providing women with pain coping skills and knowledge to advocate for themselves. The few women who were attended by a Centering facilitator valued this continuity highly. |
| Magriples U, Boynton MH, Kershaw TS, Lewis J, Rising SS, Tobin JN, Epel E, Ickovics JR. The impact of group prenatal care on pregnancy and postpartum weight trajectories. *American journal of obstetrics and gynecology*. 2015 Nov 1;213(5):688-e1. | USA | Secondary analysis of a cluster RCT; medical record review and structured interviews to evaluate the weight change trajectories in the control and intervention groups. They note lack of evidence of benefit from direct interventions and hypothesise that the group model may enhance weight management through discussion, skills building and stress reduction. |
| McDonald SD, Sword W, Eryuzlu LN, Neupane B, Beyene J, Biringer AB. Why are half of women interested in participating in group prenatal care?. *Maternal and child health journal*. 2016 Jan;20:97-105. | Canada | Self-administered questionnaire. Characteristics of women who accept to participate in gANC and why they do. Refers to existing evidence on outcomes with no further information. |
| Patil CL, Klima CS, Leshabari SC, Steffen AD, Pauls H, McGown M, Norr KF. Randomized controlled pilot of a group antenatal care model and the sociodemographic factors associated with pregnancy- | Malawi and Tanzania | Pilot RCT of Group ANC to test hypothesis that the model increases empowerment of pregnant women. Variable findings with indication that empowerment was increased for some groups (Muslim women) and settings (Malawi), but not all. Empowerment was conceptualised as ability of individuals to 'improve capacities, to critically evaluate situations and to take actions to |
| related empowerment in sub-Saharan Africa. BMC pregnancy and childbirth. 2017 Nov;17(2):1-0.<br>Patil CL, Klima CS, Steffen AD, Leshabari SC, Pauls H, Norr KF. Implementation challenges and outcomes of a randomized controlled pilot study of a group prenatal care model in Malawi and Tanzania. International Journal of Gynecology & Obstetrics. 2017 Dec;139(3):290-6. |  | improve those situations’ and understood as positively associated with uptake of maternity and reproductive services and improved infant outcomes. Empowerment was also seen as related to sense of control and ability to adopt healthy behaviours. The concept was explicitly drawn from Bandura’s self-efficacy theory, feminist and social theory and seen as supported by community building, continuity of carer and building self-care skills in gANC.<br>Linked study report focused on implementation challenges and pilot trial outcomes. |
| Riggs E, Muyeen S, Brown S, Dawson W, Petschel P, Tardiff W, Norman F, Vanpraag D, Szwarc J, Yelland J. Cultural safety and belonging for refugee background women attending group pregnancy care: an Australian qualitative study. Birth. 2017 Jun;44(2):145-52. | Australia | ‘Women reported feeling empowered and confident through learning about pregnancy and childbirth in the group setting. The collective sharing of stories in the facilitated environment allowed women to feel prepared, confident and reassured, with the greatest benefits coming from storytelling with peers, and developing trusting relationships with a team of professionals, with whom women were able to communicate in their own language. Women also discussed the pivotal role of the bicultural worker in the multidisciplinary care team.’ |
| Rijnders M, Jans S, Aalhuizen I, Detmar S, Crone M. Women-centered care: Implementation of CenteringPregnancy® in The Netherlands. Birth. 2019 Sep;46(3):450-60. | The Netherlands | Retrospective cohort study (n = 2318) and survey to investigate outcome differences between CP and traditional individual prenatal care. Focused on feasibility of implementation but refers to empowerment and satisfaction with care in a context of low satisfaction among Dutch women. |
| Sayinzoga F, Lundeen T, Gakwerere M, Manzi E, Nsaba YD, Umuziga MP, Kalisa IR, Musange SF, Walker D. Use of a facilitated group process to design and implement a group antenatal and postnatal care program in Rwanda. Journal of Midwifery & Women's Health. 2018 Sep;63(5):593-601. | Rwanda | Describes the process of adapting a Centering-based group model to the Rwandan context and development of an implementation plan. Refers to existing evidence on potential benefits. Peer (equal) approach to the group was seen as a 1<sup>st</sup> principle. No further information is provided on the potential mechanisms or principles. The article is useful primarily in setting out a consultation process for adaptation and implementation. |
| Tilden EL, Emeis CL, Caughey AB, Weinstein SR, Futernick SB, Lee CS. The influence of group versus individual prenatal care on phase of labor at hospital admission. Journal of Midwifery & Women's Health. 2016 Jul;61(4):427-34. | USA | Retrospective case control study of influence of Group Versus Individual Prenatal Care on Phase of Labor at Hospital Admission. Hypothesis that this may reduce admission in latent phase of labour as a result of improved information and understanding about labour. Significant differences were found with women who received GANC more likely to be admitted in active labour. |
| Trotman G, Chhatre G, Darolia R, Tefera E, Damle L, Gomez-Lobo V. The effect of centering pregnancy versus traditional prenatal care models on improved adolescent health behaviors in the perinatal period. Journal of pediatric and adolescent gynecology. 2015 | USA | Known features of this model - education, group discussion, and social support considered helpful to address needs of adolescents for support, to reduce isolation and increase healthy behaviours. Improved compliance with prenatal visits, uptake of LARC methods, adequate weight gain, and increased rates of breastfeeding were identified and seen as associated with improved healthy habits via psychosocial support and education. |
| Oct 1;28(5):395-401. |  |  |
| Tubay AT, Mansalis KA, Simpson MJ, Armitage NH, Briscoe G, Potts V. The effects of group prenatal care on infant birthweight and maternal well-being: a randomized controlled trial. Military medicine. 2019 May 1;184(5-6):e440-6. | USA | RCT with women from the military receiving either traditional one-on-one prenatal care or group prenatal care. Focused on clinical outcomes and satisfaction with care. No differences were found apart from improvement in births at appropriate gestational age. Refers to existing theories of the mechanism of effect as peer support and self-management strategies that mitigate stress and reduce high-risk behaviours, and also through reducing isolation, which they see as particularly relevant for a military population. |
| Walton RB, Shaffer S, Heaton J. Group prenatal care outcomes in a military population: a retrospective cohort study. Military medicine. 2015 Jul 1;180(7):825-9. | USA | Retrospective Cohort Study of outcomes in a military population. No significant differences in outcomes were found, which they attributed to small study size. Centering Pregnancy referred to as an integrative approach, combining education, peer support, and family members that has been associated with improved outcomes in some studies. No further information given on the theory. |
| Yorga KD, Sheeder JL. Which pregnant adolescents would be interested in group-based care, and why?. Journal of Pediatric and Adolescent Gynecology. 2015 Dec 1;28(6):508-15. | USA | Study focused on factors that influence pregnant adolescents' interest in participating in gANC or not. Key reasons to participate were identified as to belong to a peer group, to receive additional education and support and to have fun. Structured survey including some open-response questions. Hypotheses discussed were that the approach may reduce loneliness and isolation for pregnant adolescents, and that it may provide an environment of positive peer support which also supports increase in knowledge, which in turn may enhance healthy behaviours. |
| Zorrilla CD, Sánchez I, Mosquera AM, Sierra D, Pérez LA, Rabionet S, Rivera-Viñas J. Improved infant outcomes with group prenatal care in Puerto Rico. Source journal of obstetrics and gynaecology. 2017;1(1). | Puerto Rico | Retrospective chart review looking at outcomes in Puerto Rican population. Improvements were found in birthweight and gestational age. No information given about rationale for care except that the model has been found to improve uptake and outcomes in other settings. |

